# Paving the way for precision treatment of psychiatric symptoms with functional connectivity neurofeedback

**DOI:** 10.1101/2024.04.03.24304187

**Authors:** JE Taylor, T Oka, M Murakami, T Motegi, T Yamada, T Kawashima, Y Kobayashi, Y Yoshihara, J Miyata, T Murai, M Kawato, A Cortese

## Abstract

**Introduction:** Major depressive disorder (MDD) remains challenging to treat, with many patients failing to respond adequately to existing therapies. Patients with MDD have heterogeneous subsets of symptoms with differing underlying neural aberrations. Treatment response may improve if treatments become more individualised. We recently showed preliminary evidence that normalisation of a neural network and a corresponding reduction in related symptoms can be achieved using a Brain Machine Interface (BMI) called real-time fMRI functional connectivity neurofeedback (FCNef). However, the robustness of this effect, and the best FCNef parameters for optimising therapeutic outcomes remained unknown.

**Methods:** We ran additional participants, with a final dataset of N = 68, in our FCNef protocol. Functional connectivity between the dorsolateral prefrontal cortex (DLPFC) and posterior cingulate cortex/precuneus (PCC) was targeted with the goal of reducing brooding rumination symptoms. Core FCNef parameters (experimental schedule and reward) were manipulated between participants.

**Results:** We replicated findings that normalisation of DLPFC-PCC connectivity with FCNef correlates significantly with reductions in brooding rumination, but not with changes in anxiety, which is associated with different neural circuits. The difference between these correlations was significant, highlighting the precision of this effect. Finally, we found that successful DLPFC-PCC normalisation and corresponding changes in brooding rumination depended on specific FCNef parameters. The most effective protocol involved consecutive training days with greater external reward.

**Conclusions:** These results highlight the potential of FCNef for precision medicine in psychiatry and underscore the importance of optimising parameters to enhance feasibility of BMI-based clinical interventions.

## 1. Introduction

The World Health Organization estimates that major depressive disorders will become the top cause of global disease burden by 2030 [1]. Of patients who do receive treatment, an estimated 30-50% do not respond fully [2,3]. Patients with the same clinical diagnosis can have heterogeneous subsets of symptoms that relate to different underlying neural mechanisms [4]. Nonetheless, they usually receive relatively homogenous treatment. For example, all or most clinical practice guidelines recommend selective serotonin reuptake inhibitors as first-line treatment for depression [5]. To improve response rates, individual differences clearly need to be considered. Future treatment may become more individualised using Brain-Machine Interfaces (BMIs) to identify and target underlying patient neural aberrations. However, medical regulatory approval presents serious challenges to this [6]. To date, only a handful of BMIs have received approval from local medical regulatory agencies for human trials [7–9], and even fewer have received full market authorisation [10,11]. A key step toward approval of a given BMI technique by regulatory agencies is demonstrating the optimality of chosen parameters. Here, we extend previous results to better examine the robustness of a promising form of BMI called Functional-Connectivity Neurofeedback (FCNef) [12], and to systematically investigate a specific set of its parameters.

FCNef is a functional magnetic resonance imaging (fMRI) neurofeedback technique in which participants receive real-time feedback about the current state of functional connectivity between targeted brain regions (measured as the correlation between time-courses of BOLD activity from these regions). This feedback is used to train participants to make a targeted functional connection more positive or negative, a result that has been demonstrated in multiple studies [12–19]. Showing promise for precision medicine, recent FCNef studies have reported precise correspondence between normalisation of functional connections and reductions in specifically related symptoms [12,17,18,20]. Consistent with these results, we previously found that normalisation of functional connectivity between the dorsolateral prefrontal cortex (DLPFC) and the precuneus/posterior cingulate cortex (PCC) that occurred with FCNef was related to reductions in brooding rumination, but not anxiety symptoms. Importantly, these effects persisted at least 1-2 months after FCNef [12]. Nonetheless these results were reported from only a small sample. We have since continued to collect data and here we examine the robustness of these effects by testing for their replication in the newly collected data. Furthermore, because combining the new and old data provides sufficient statistical power to properly compare correlation coefficients, for the first time, we are able to directly examine the specificity of this effect.

Given its potential use as a medical tool, FCNef should maximise health outcomes while minimising patient burden. Here, we operationalised FCNef success as the normalisation of DLPFC/PCC functional connectivity and a related reduction in brooding (but not anxiety) symptoms. We sought to find parameters that would best enhance this, while also keeping participant fatigue to a minimum (because preliminary testing in a clinical sample caused fatigue-related drop-out of one out of six patients [21]). To accomplish our objective, we focused on the following parameters: (1) Reward schedule: During real-time neurofeedback tasks, feedback has conventionally been provided simply as scores that reflect how similar the induced brain activity is to the target brain activity [22–24]. However, recent evidence suggests that target neural activity may be better reinforced during neurofeedback when external reward, such as money, is also used [25]. Here, we manipulated how bonus money was assigned to feedback scores so that different groups of participants could earn less/more overall external reward. (2) Experimental schedule. Most FCNef studies [15], including our own [12,19], have required participants to come in for multiple consecutive days of experimentation. This can be exhausting and requires motivation and organisation skills that can be diminished in psychiatric disorders [26]. Therefore, we tested whether a more flexible schedule, over non-consecutive days, could yield similar results to the consecutive training schedule.

Overall, we ran 68 participants in our FCNef for depression paradigm while manipulating reward schedule (low/high) and experimental schedule (consecutive/non-consecutive training days). Our goals were twofold: (1) using a larger sample size to examine the robustness of our previously reported results and more precisely examine the specificity of the FCNef effect, and (2) to fine-tune underlying parameters.

## 2. Methods

### 2.1. Participants

Participants were screened twice using the Beck Depression Inventory-II (BDI) questionnaire [27] (see Supplementary Methods for more detail). Only those with an average score ≥ 8, who indicated no intention of committing suicide, who spoke Japanese, who held no current clinical diagnosis, and who were not currently receiving treatment for a psychiatric illness were invited to participate. Overall, 69 people passed the screening and participated in the main experiment. However, experimental data of one participant was subsequently excluded from data analysis because it was revealed, subsequent to experimentation, that that subject held a current clinical diagnosis. Of the 68 participants whose data were used for analyses, the average BDI score over the two screening measurements was 14.33, with a standard deviation (STD) of 5.26. This puts them generally in the category of “mild depression” [27]. For this reason and because these participants lacked current clinical diagnoses, we considered them “subclinical”.

### 2.2. Experimental Conditions

We wished to examine the overall success of our paradigm and to determine whether it is influenced by reward and experimental schedules. However, due to financial and time constraints (it takes 10 days of experimentation to screen and run just one participant in this paradigm), we could only run participants in three experimental groups: those with consecutive days of experimentation and a high-reward schedule (hereafter, “Consec/High-Rew”), those with consecutive days of experimentation and a low-reward schedule (hereafter, “Consec/Low-Rew”), and those with non-consecutive days of experimentation and a low-reward schedule (hereafter, “Non-Consec/Low-Rew”) (see Table 1 for sample size details). Participants in the three groups did not differ significantly in baseline levels of our main measures of interest: (1) DLPFC-PCC resting-state functional-connectivity (rs-FC), (2) BDI scores, and (3) brooding scores (measured on a subscale of the Rumination Response Scale [28,29]) (see Supplementary Table 7). However, we found that baseline anxiety levels (measured with the trait anxiety subscale of the State-Trait Anxiety Inventory [30]) differed significantly between the Consec/Low-Rew and Non-Consec/Low-Rew groups (see Supplementary Table 7). This is unlikely to have had a large impact on results because: (a) the rs-FC we targeted is not thought to relate to anxiety, and (b) results showed that differences in FCNef success were not greatest between these two groups (as can be seen further down).

**Table 1.** Sample sizes for three groups of participants. *This table shows the sample sizes for: (1) participants run in consecutive days of FCNef with the high-reward schedule (the “Consec/High-Rew group”), (2) those run in consecutive days of FCNef with the low-reward schedule (the “Consec/Low-Rew group”), and (3) those run over non-consecutive days of FCNef with the low-reward schedule (the “Non-Consec/Low-Rew group”). No participants were run over non-consecutive days of FCNef with the high-reward schedule (no “Non-Consec/High-Rew” group). Sample sizes are shown for the main experiment and for one- and two-month follow-up tests. There were fewer participants for the follow-up tests for the Consec/High-Rew group because the long-term tests were not included in the earliest stages of this experiment, so only 12 participants from this group were invited back*.

| Group | Main Experiment<br>Sample Size | 1-Month Follow-up Sample Size | 2-Month Follow-up Sample Size |
| --- | --- | --- | --- |
| Consec/High-Rew | 21 | 11 | 9 |
| Consec/Low-Rew | 23 | 20 | 17 |
| Non/Low-Rew | 24 | 22 | 20 |
| Total | 68 | 53 | 46 |

Participant group assignments were determined by concurrent availability of participants, experimenters, and MR machine facilities. This allocation method reflects real-world constraints in neuroimaging research, though we acknowledge it could have introduced potential self-selection effects. To address possible allocation bias, we conducted analyses to compare baseline demographic measurements between groups (age, sex): No meaningful differences were found (see Supplementary Results). It should be noted that data of 19 participants (9/21 from the Consec/High-Rew group and 10/23 from the Consec/Low-Rew group) have been reported elsewhere [12]. Further details about recruitment, participant demographics, and payment can be found in the Supplementary Methods.

### 2.3. Experimental procedure, materials, and imaging data acquisition

An outline of the experimental procedure is shown in Figure 1 alongside details of the FCNef task. These were largely the same as in our previous report [12], except for specific experimental conditions of interest. These experimental conditions are described in the sub-section below entitled “Differences in experimental conditions.” The protocol and imaging data acquisition details are identical to those in our previous report [12] and are summarised in the Supplementary Methods.

**Figure 1.**
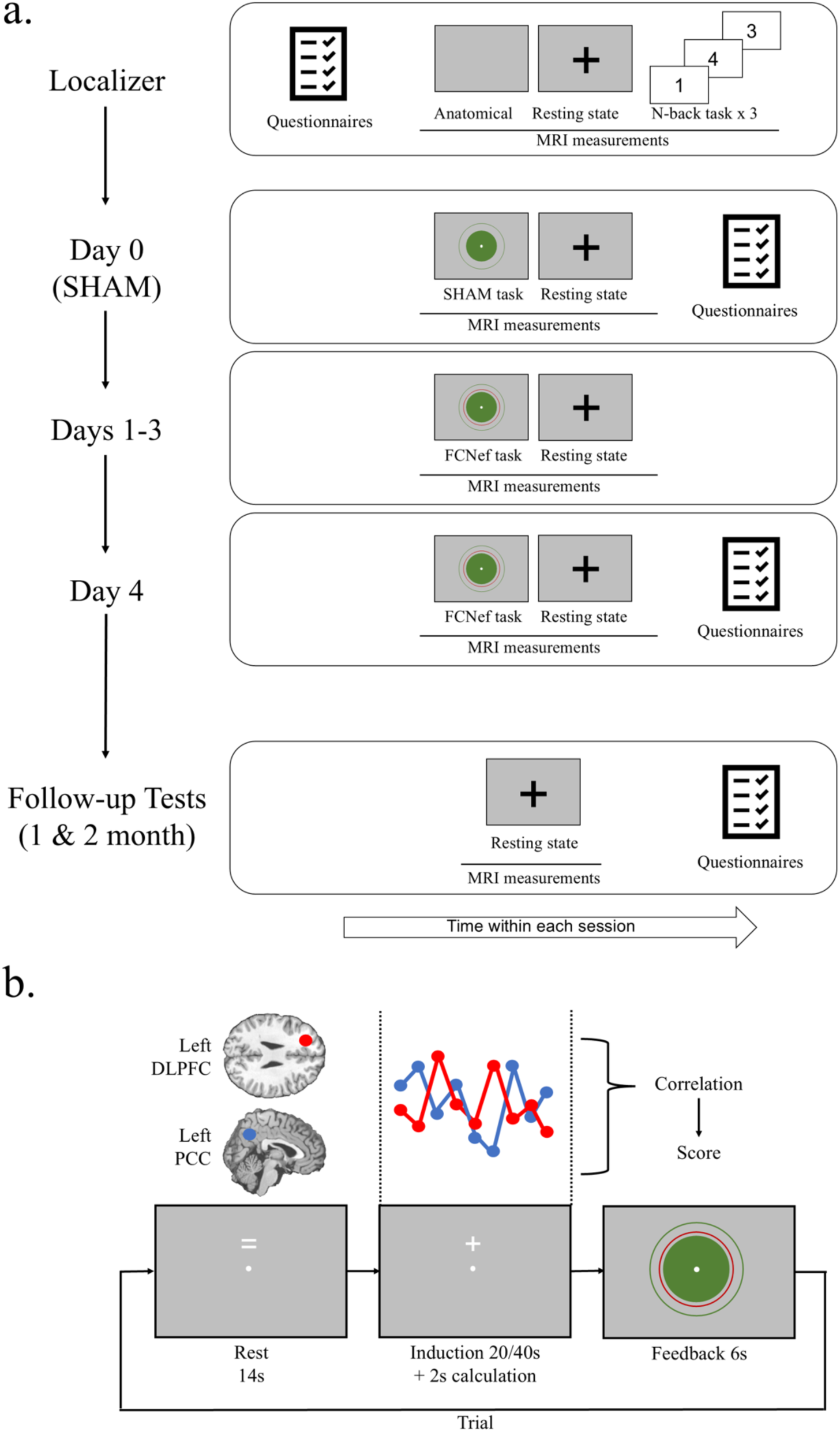
Experimental procedure and example FCNef trial. **a. *The order of events on each day of experimentation.*** *Questionnaires = the Beck Depression Inventory-II* [27]*, the Rumination Response Scale* [28]*, and the State-Trait Anxiety Inventory* [30]*. Anatomical = T1-weighted structural MRI. N-back = a well-known executive control task* [54]*, used here as a functional localiser*[12]*. **b. An example FCNef trial.** During the rest period, participants were to simply relax. During the induction period, they were asked to “somehow” manipulate their brain activity to get the best possible feedback. Participants were told that different strategies of brain activity manipulation might work for different people. Unbeknown to participants (nothing changed on screen), there was a 2s calculation period at the end of the induction period. During FCNef, DLPFC-PCC connectivity (from the induction period) was calculated during the calculation period and this determined the feedback presented during the feedback period. During SHAM, however, feedback was just random. Feedback was presented on screen as a green circle and participants had been clearly instructed that the larger this was, the more monetary reward they would receive on that trial. During FCNef, they were instructed to try to make the green circle bigger than the red circle that was also presented on screen. The circumference of this red circle represented the participant’s baseline DLPFC-PCC connectivity (the average from SHAM). During SHAM, there was no red circle and participants were simply instructed to try to make the green circle as big as possible. Modified with permission from Taylor et al. (2022)*[12].

Details of the symptom questionnaires can be found in the Supplementary Methods, but overall general depressive symptoms were measured with the BDI [27]. Brooding rumination symptoms were measured with a subscale of the Rumination Response Scale (RRS) [28,29], and trait anxiety symptoms were measured with the trait anxiety subscale of the State-Trait Anxiety Inventory (STAI-Y2) [30]. As can be seen in Figure 1, because there would be limited clinical meaning, these symptom questionnaire scores were not measured on all days of the main experiment. Instead, they were only measured on the first day (Day 0) and last day (Day 4) of the main experiment and during one- and two-month follow-up tests.

### 2.4. Differences in experimental conditions

The following parameters were manipulated, such that some participants were run under conditions different from those reported in our previous papers [12,19].

#### 2.4.1. Reward schedule

All participants received a baseline reward bonus of ¥500 on each day of the SHAM and FCNef sessions. This is the maximum reward they could receive in the SHAM task, but they could get an additional reward bonus in the FCNef task depending on their average FCNef scores from that day.

Importantly, two calculation methods were used to determine this additional reward bonus. Participants in the low-reward schedule conditions could achieve an additional reward bonus of ¥100 for each average FCNef score point over 75. Instead, participants in the high-reward schedule condition could achieve an additional reward bonus of ¥50 for each average FCNef score point over 50. See Supplementary Table 2 for specific examples. The way in which scores were calculated in the SHAM and FCNef tasks was identical across conditions. This was the same method as in our previous report [12] and is described in the Supplementary Methods.

#### 2.4.2. Experimental schedule

Two groups of participants completed Days 0-4 over 5 consecutive days (Monday-Friday). The third group completed Days 0-4 over 5 non-consecutive days, which took place over a period of several weeks to months (mean of 18.5 days ± STD of 15.3 days).

#### 2.4.3. Induction time-window

Consistent with earlier versions of our FCNef for depression paradigm [12,19], about half of the participants from each group completed FCNef and SHAM with a 40 sec induction time-window (see Supplementary Table 1). Because re-analysis of pilot data suggested it would not affect results (see Supplementary Results, Supplementary Table 3, and Supplementary Figure 1), the other half of participants completed FCNef and SHAM with a 20 sec induction time-window. We included this manipulation with the goal of improving our paradigm. Using a 20 sec time-window reduces the overall experiment by about 10 mins each day, which could be crucial for patients who tire easily. Indeed, when we checked the data of our three groups of participants, we found that our measure of FCNef success did not change in any meaningful way that depended on the induction time-window (see Supplementary Results and Supplementary Tables 4-6). Therefore, in the main text of this report, we did not split the data by induction time-window because (a) it is hard to draw strong conclusions about null effects, and (b) splitting the data further like this would make sample sizes in each cell even smaller, making it hard to draw conclusions about other parameters of interest.

### 2.5. Data analyses

#### 2.5.1. Correlations to extend previously reported results

We previously reported correlations between rs-FC and symptom changes using data from 19 participants who were run over consecutive days of FCNef [12]. Ten of these participants were run with the high-reward schedule and the other nine were run with the low-reward schedule, but overall their combined data showed promising results. Since then, we have collected data from 25 more participants under the same experimental conditions (consecutive days of FCNef with low/high-reward schedules), which gives us an overall dataset with 44 participants run under these conditions (see Table 1). Note that this does not include the data of the Non-Consec/Low-Rew group, which has also been collected since our previous report. This is because this group was run with a different experimental parameter to our previous report (non-consecutive days of experimentation) and so its data cannot contribute directly to our replication test. One goal of collecting this additional data was to assess the robustness of previously reported results in a larger dataset. Before doing this in the 44 participants, however, we first re-ran relevant correlations using only data of the additional 25 participants to ensure that we were not introducing bias by adding new data to the existing pool. After this test, we then re-ran the previously reported correlations with the larger pooled dataset. To check the adequacy of the sample size for statistical tests, we conducted post-hoc power analyses with G*Power version 3.1.9.7 (Franz Faul, Kiel University, Germany). The significance threshold was set at *α* = 0.05, and we input relevant correlational values to calculate statistical power.

#### 2.5.2. Linear Mixed Effect (LME) models to examine parameters of interest

We ran multiple LME models to examine how multiple measures related to FCNef success were impacted by our manipulated parameters (see Table 2). Dependent variables (DVs) in these models were taken from the task (e.g. average task score) or from the resting-state scans (e.g. changes in resting-state functional connectivity from before to after FCNef). All 3 groups of participants (N = 68) were included in these analyses so we could best examine parameters of interest.

**Table 2.**
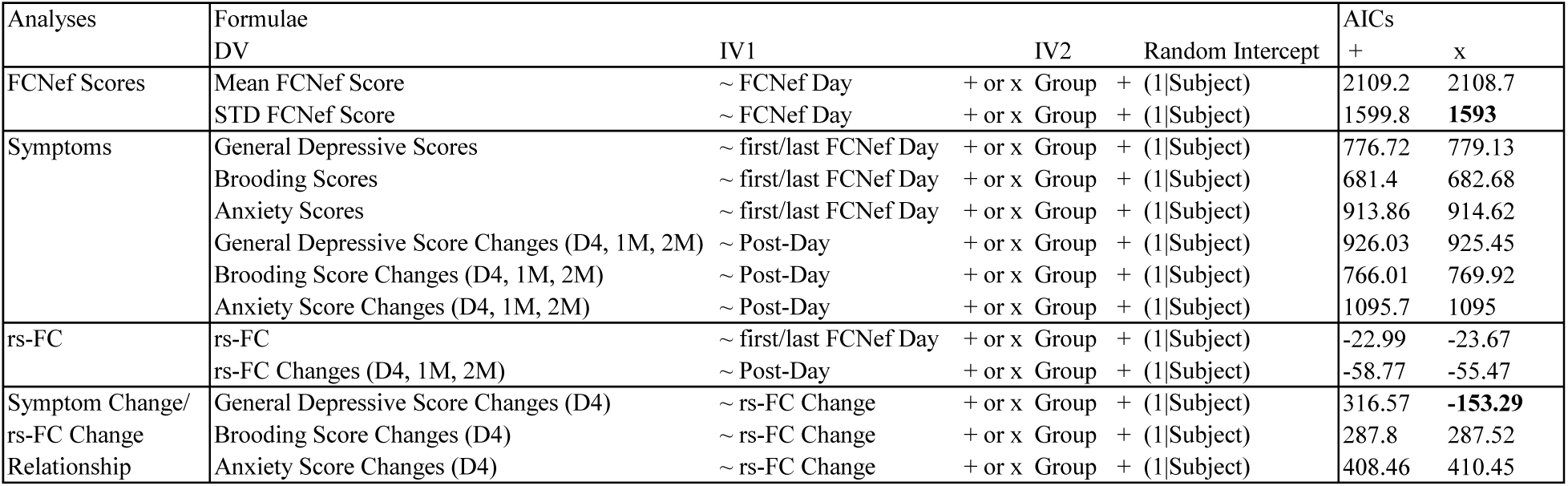
The models used to examine dependent variables from the FCNef experiment. *Models with and without interactions between independent variables (IVs) were compared to see which would best predict each dependent variable (DV). Akaike Information Criteria (AIC) are displayed in columns labelled ‘+’ (for models without interactions) and ‘x’ (for models with interactions). These are highlighted in bold in cases in which likelihood ratio testing showed the corresponding model to be significantly better fit than the alternative model. If there was no significant difference, then the model without the interaction was selected as best-fit because it was the simplest. FCNef Day = Days 1-4; Subject = experimental participant; STD = standard deviation; First/last FCNef Day = Days 0 and 4; Post-Day = Day 4, and the 1- and 2-month follow-up test days; Changes = data from the day indicated in brackets (e.g. D4) minus data from Day 0; rs-FC = resting state functional connectivity between the DLPFC-PCC*.

We were unable to run a Non/High-Rew group (see Table 1). This means we could not run full factorial analyses to investigate all possible interactions between parameters that we manipulated, although we had no reason to expect interactions anyway. Instead, in all models reported here, we included “Group” as a categorical Independent Variable (IV) with 3 levels. By nature, this type of analysis compares the variance related to the second two levels with the variance related to the first level (the reference), so it is important to consider what data will be input at what level. Here, we decided to set the Consec/High-Rew group as the first level because this group had parameters that we used in the initial pilot experiment [19] and we wanted to examine whether changes to these initial parameters would improve or worsen FCNef success.

In addition to Group, the LME models also included other relevant IVs, which differed between models depending on the DV. In models examining how brain changes predicted symptom changes, this was changes in resting-state functional connectivity from before to after FCNef. In other models, this was the relevant experimental days.

For each DV, to explore the data, we ran LME models with and without the interaction term between IVs. We then tested these against one another to see which best fit the data.

No interaction term: *‘DV∼X+Group+(1|Subject)’*

With interaction term: *‘DV∼X*Group+(1|Subject)’*

Likelihood ratio tests were used to compare models with and without interaction terms for all DVs. If there was a significant difference between models, the best-fit model was always selected based on the lowest Akaike Information Criterion (AIC, a measure of goodness of fit, which can be used to assess a model’s relative quality [31]). If there was no significant difference between models then the simplest model, that without interaction, was selected as best-fit. Detailed information about the best-fit of each of these pairs of models can be found in tables in the Supplementary Results (Supplementary Tables 8, 9, 11-17, 19-24), with all significant main effects and interactions from the best-fit models additionally being reported in the Results Section below.

#### 2.5.3. Follow-up statistical testing

Significant main effects and interactions from the best-fit models, as well as effects that we had specific hypotheses about, were followed up with t-tests or correlations. We applied False Discovery Rate (FDR) correction [32] whenever there were multiple comparisons. We had strong directional hypotheses that symptoms would decrease from before to after FCNef and that rs-FC would become more negative, aligning with patterns observed in healthy individuals. Therefore, we conducted the related t-tests using a one-tailed approach. Similarly, based on our strong directional hypothesis that symptoms would decrease as rs-FC became more negative (indicating a positive correlation between changes in symptoms and changes in rs-FC), we also used a one-tailed approach for the related correlation analyses.

## 3. Results

### 3.1. Extension of past results

#### 3.1.1. Previously reported data

In our previous publication [12], we reported correlations between rs-FC and symptom changes using combined data of 9 participants from the current Consec/High-Rew group and 10 participants from the current Consec/Low-Rew group. We found a significant positive relationship between general depression change and rs-FC change (*r* = 0.78, *p_FDR_* = 0.0001, 95% CI [0.51, 0.91]) and between brooding change and rs-FC change (*r* = 0.43, *p_FDR_* = 0.048, 95% CI [-0.27, 0.74]), but a non-significant negative relationship between anxiety change and rs-FC change (*r* = 0.11, *p_FDR_* = 0.32, 95% CI [-0.36, 0.54]) (note that we previously only reported correlation coefficients and uncorrected *p*-values). Because we specifically hypothesised that changes in DLPFC-PCC rs-FC from before to after FCNef should relate to changes in brooding but not anxiety symptoms, here we calculated Z-tests to compare these correlation coefficients [33]. These were run with one tail due to our directional hypothesis, and no significant difference was found (*z* = 0.88, *p* = 0.16). A post-hoc power analysis was conducted using correlation coefficients used for this Z-test (*r* = 0.43 and *r* = 0.11) and the coefficient for the correlation between brooding and anxiety changes (*r* = -0.03). This revealed a post-hoc power of 0.25, suggesting that the sample size (N = 19) was statistically insufficient to detect an effect of this magnitude.

#### 3.1.2. Newly collected data

We have since collected more data for each of these groups (12 more participants for the Consec/High-Rew group and 13 more participants for the Consec/Low-Rew group) with the goal of testing robustness of the aforementioned findings in a larger dataset. To ensure that we were not adding bias or overestimating effects, before testing results for the pooled larger dataset, we first ran a replication test using just the newly collected data (N = 25). Consistent with the aforementioned results, we found a positive relationship between general depression change and rs-FC change (*r* = 0.26, *p_FDR_* = 0.16, 95% CI [-0.16, 0.60]) and between brooding change and rs-FC change (*r* = 0.40, *p_FDR_* = 0.08, 95% CI [-0.01, 0.69]), although these did not reach significance here. We found a non-significant negative relationship between anxiety change and rs-FC change (*r* = -0.17, *p_FDR_* = 0.80, 95% CI [-0.53, 0.24]).

#### 3.1.3. Pooled data

Overall, we consider the previously reported effects to be replicated in the newly collected data because confidence intervals overlapped and effect sizes were comparable. When we next pooled these data (total N = 44) we found a significant correlation between general depression change and rs-FC change (*r* = 0.48, *p_FDR_* = 0.002, 95% CI [0.21, 0.68]), and between brooding change and rs-FC change (*r* = 0.42, *p_FDR_* = 0.004; 95% CI [0.13, 0.64]), but not between anxiety change and rs-FC change (*r* = -0.03, *p_FDR_* = 0.57; 95% CI [-0.32, 0.27]). Demonstrating the specificity achieved when targeting the DLPFC/PCC functional connection, we found a significant difference between coefficients from brooding change/rs-FC change and anxiety change/rs-FC change correlations (*z* = 2.15, *p* = 0.016) (see Figure 2). A post-hoc power analysis was conducted using correlation coefficients employed in this Z-test (*r* = 0.42 and *r* = -0.03) and the coefficient for the correlation between brooding and anxiety changes (*r* = 0.23).

**Figure 2.**
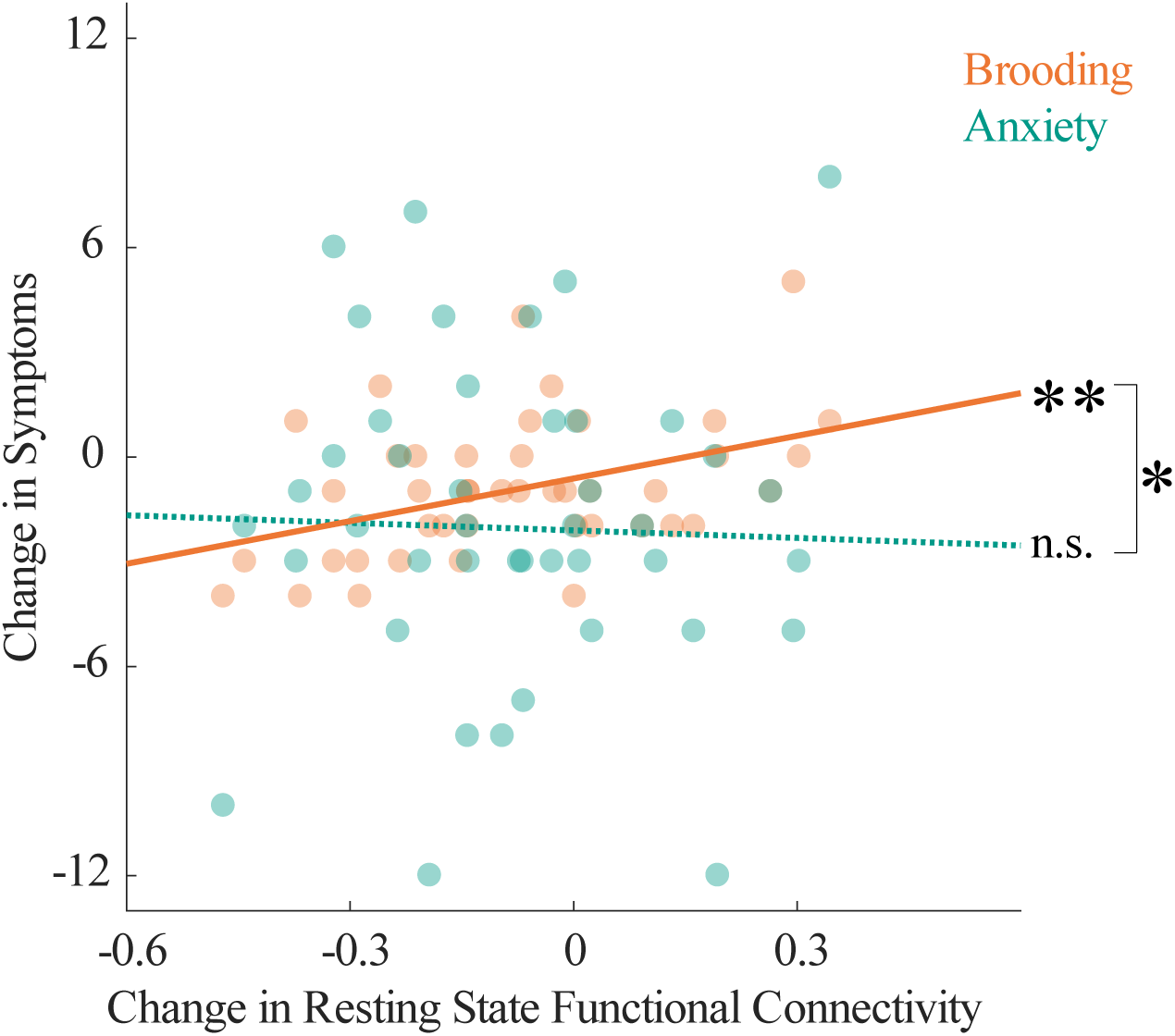
Comparison of rs-FC/Brooding and rs-FC/anxiety correlations. *Promising for precision medicine, changes in DLPFC-PCC resting state functional connectivity (rs-FC) from before to after FCNef were significantly correlated with changes in brooding, but not anxiety symptoms. Pooling participants from consecutive conditions (to be consistent with conditions used in our previous report) provided us with the statistical power to compare these correlation coefficients. The significant difference found, highlights the precision of FCNef targeting this functional connection. Note that this same data, but split by condition, is presented again in* Figure 5a *and 5b. ** = p_FDR_* < 0.01, * = *p* < 0.05.

This revealed a power of 0.79, suggesting that the sample size (N = 44) was sufficient to detect an effect of this magnitude with high probability.

### 3.2. Parameter Investigations

#### 3.2.1. FCNef scores

In the total dataset (N = 68), while comparing groups, we first examined whether mean FCNef scores increased significantly across training days, because this would indicate that participants successfully modified the target functional connection in the trained direction. Additionally, we assessed whether variance in FCNef scores decreased significantly across training days, because this would indicate that participants had gained better control of the DLPFC-PCC functional connection.

The best-fit LME model to predict means of average daily FCNef scores was the model with no interaction (Table 2 and Supplementary Table 8). This model showed a significant main effect of FCNef day (*p* = 0.0009). In short, mean scores increased over training days (see Figure 3), which indicates that participants could do the task. There was also a significant effect of the Non-Consec/Low-Rew group (*p* = 0.046): Overall scores were lower for this group than for the reference Consec/High-Rew group (see Figure 3a), who were the only group capable of raising their scores significantly above baseline.

**Figure 3.**
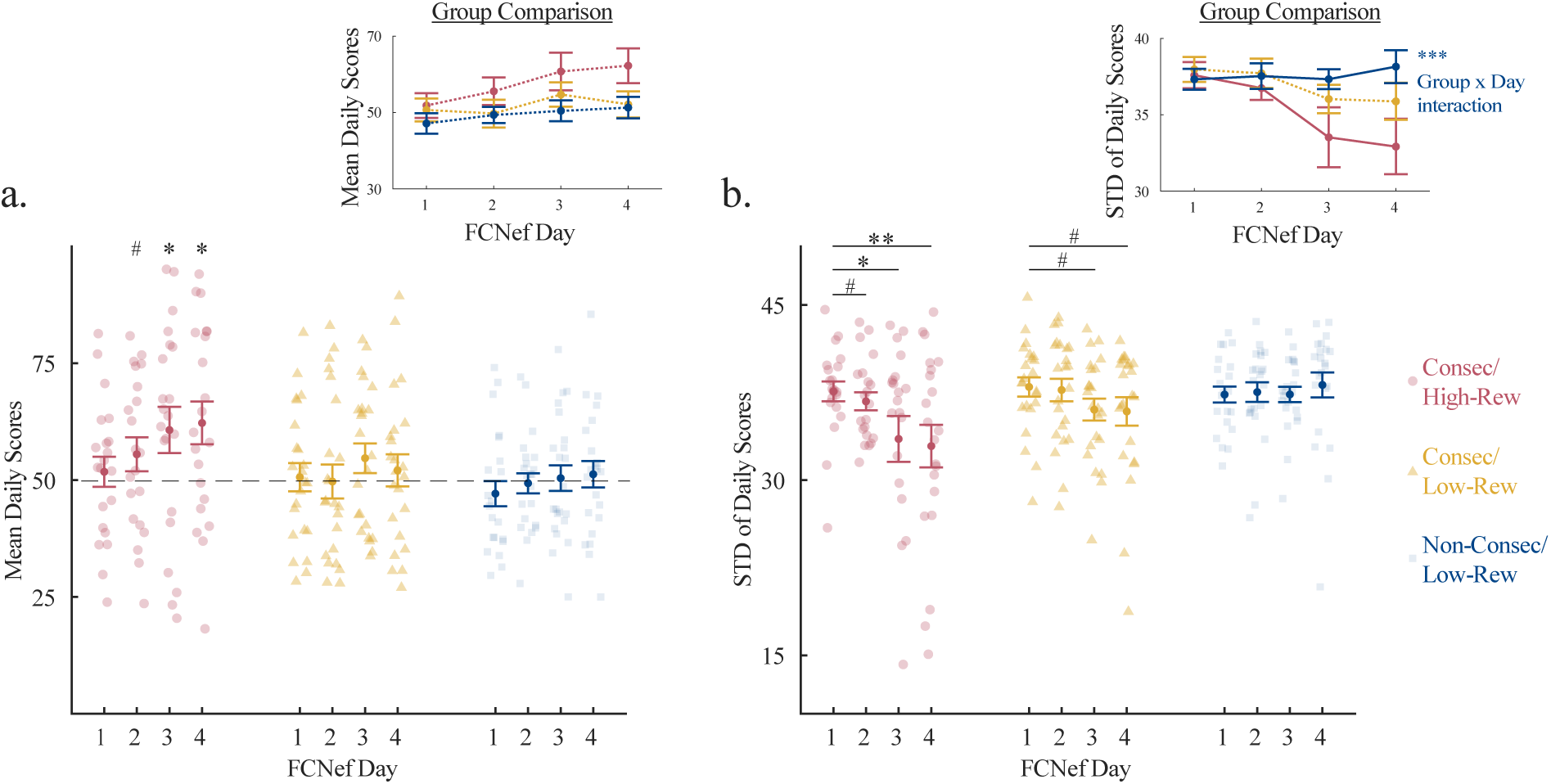
Means and Standard Deviations of FCNef scores. **(a)** *Mean daily FCNef scores. Investigative t-tests were conducted separately for each group to examine whether scores on each FCNef day were higher than a baseline score of 50 (which was the average score given during SHAM on Day 0). Scores improved significantly over FCNef days only for the Consec/High-Rew group. The Group Comparison inset shows the mean ± standard error of mean daily scores overlaid for the different groups. **(b)** Daily STDs in FCNef scores. Investigative t-tests were conducted separately for each group to examine whether variance in scores on each subsequent day of FCNef was lower than that on the Day 1 of FCNef. Score variance decreased significantly over FCNef days only for the Consec/High-Rew group, but a similar trend was found for the Consec/Low-Rew group. The Group Comparison inset shows the mean ± standard error of STD daily scores overlaid for the different groups; This highlights the Group x Day interaction found in the best-fit model for the Non-Consec/Low-Rew group relative to the reference Consec/High-Rew group. *** represents p = 0.001, ** represents pFDR < 0.01, * represents pFDR < 0.05, # represents pFDR < 0.1, STD = standard deviation*.

The best-fit model to explain variance in FCNef scores did include the interaction term (see Table 2 and Supplementary Table 9). This model showed a significant main effect of FCNef day (*p* < 0.0001), which was qualified by a significant interaction between the Non-Consec/Low-Rew group and FCNef day (*p* = 0.001). This interaction indicates that changes in FCNef score variance over FCNef days did not follow the same trajectory for the Non-Consec/Low-Rew group as it did for the reference Consec/Low-Rew group; As can be seen in Figure 3b-the variance decreased over days for the reference group, but not for the Non-Consec/Low-Rew group. No such significant interaction was found between the Consec/Low-Rew group and FCNef Day (*p* = 0.11), which indicates that the Consec/Low-Rew group’s trajectory did not differ from that of the reference group. Indeed, the data for the Consec/Low-Rew group lay between that of the other two groups, showing a trend for a decrease in variance over days (see Figure 3b).

#### 3.2.2. Self-report symptom scores

We analysed symptom questionnaire scores and compared these between the three groups as a secondary measure related to FCNef effects (Supplementary Table 10). We did not include symptom changes from before to after FCNef and into the long-term in our operationalisation of FCNef success because symptom reduction could occur for multiple reasons, including the placebo effect. Symptom reduction would be meaningful here only if specifically related to changes in the targeted brain activity.

Nonetheless, we monitored symptom changes to ensure they had not worsened over the course of the study. Based on preliminary results [12], we predicted that symptoms should decrease from before to after FCNef and then remain lower across post-days. Therefore, we used separate LME models to examine initial and long-term effects.

##### 3.2.2.1. Symptoms: The initial effect

For the best-fit models to predict both general depressive symptoms and brooding symptoms (see Table 2 and Supplementary Tables 11 and 12), we found main effects of first/last FCNef day (*p* < 0.0001 for general depressive symptoms, *p* = 0.0003 for brooding symptoms), but no main effects of group (*ps* > 0.05). Follow-up t-tests showed that these symptoms were lower on day 4 than on day 0 (*t*(67) = 4.78, *p_FDR_* < 0.0001 for general depressive symptoms; *t*(67) = 4.33, *p_FDR_* < 0.0001 for brooding symptoms). For the best-fit models to predict anxiety symptoms (see Table 2 and Supplementary Table 13), we found a significant main effect of first/last FCNef day (*p* = 0.003). A follow-up t-test showed that overall, anxiety symptoms were lower on day 4 than on day 0 (*t*(67) = 2.95, *p_FDR_* = 0.007). In this model there was also a significant main effect of the Non-Consec/Low-Rew group (*p* = 0.042), and a trend for a main effect of the Consec/Low-Rew group (*p* = 0.0995). Compared to the reference Consec/High-Rew group, participants in the Non-Consec/Low-Rew group had higher anxiety and participants in the Consec/Low-Rew group had lower anxiety (on Days 0 and 4) (see Supplementary Table 10). If anxiety has any effect on overall FCNef success, then we should see deviations from the reference group in opposite directions for these two groups, which we did not.

##### 3.2.2.2. Symptoms: Changes in the long-term

Next, we examined whether any post-changes in symptoms differed or remained stable over the long-term. For all symptom types, the best-fit model (see Table 2 and Supplementary Tables 14-17) had no significant main effect of post-day or group (*ps* > 0.05). This may mean that after symptoms decreased from day 0 to day 4, they remained at this new reduced level in the long-term. To examine this possibility, we used t-tests to compare symptoms from follow-up tests with symptoms from day 0. Indeed, we found that relative to symptoms from day 0, general depressive and brooding symptoms from the 1-month and 2-month follow-up tests were significantly reduced (*t*(52) = 4.28, *p_FDR_* = 0.0001 for general depressive at 1-month, *t*(52) = 4.28, *p_FDR_* < 0.0001 for brooding at 1-month, (*t*(45) = 2.24, *p_FDR_* = 0.015 for general depressive symptoms at 2-months, *t*(45) = 5.60, *p_FDR_* < 0.0001 for brooding symptoms at 2-months). We found a similar pattern, but non-significant, for anxiety symptoms (*t*(52) = 4.28, *p_FDR_* = 0.061 at 1-month, *t*(45) = 0.92, *p_FDR_* = 0.18 at 2-months). Overall, these results show that general depressive and brooding symptoms significantly decreased from before to after FCNef and remained at this new reduced level in the long-term. Anxiety symptoms also decreased significantly from before to after FCNef and remained in this same direction in the long-term, albeit not significantly. There was no persuasive evidence that group, and therefore the manipulated parameters, influenced these results.

#### 3.2.3. Resting-state functional connectivity

If FCNef is successful, then we predict that participant rs-FC should become more negative with FCNef, which was the trained direction, and that it should remain so across the post-days. Therefore, we used separate LME models to examine rs-FC initial and long-term effects for the three groups of participants. Note that we measured rs-FC on all days (0-4) of the main experiment. However, we only included data from days 0 and 4 in LME models to examine the initial effect. This is because rs-FC on days 1-3 may be subject to homeostatic and/or compensatory mechanisms, causing overshoots [12] or rebounds [34] in brain activity. Average rs-FC for each group for each day of experimentation can be seen in Supplementary Table 18.

##### 3.2.3.1. rs-FC: The initial effect

Likelihood ratio testing showed that models to predict rs-FC from days 0 and 4 did not differ significantly, depending on whether an interaction term was included. Therefore, we selected the model without the interaction term as best-fit, because it was simpler (see Table 2 and Supplementary Table 19). This model showed a trend for a main effect of first/last FCNef day (*p* = 0.094), but no main effect of group (*ps* > 0.05). Nonetheless, Figure 4 clearly shows that rs-FC on day 4 was more negative relative to baseline for the Consec/High-Rew group, but not for the Non-Consec/Low-Rew group. rs-FC changes from day 0 to day 4 for the Consec/Low-Rew group lay in between those of the other 2 groups (see Figure 4). Evidence for these between-group differences may be found at the LME model level if this experiment is run with greater sample sizes; This is suggested by the fact that the model with the interaction term trended toward being better than the model without, despite this not reaching full significance (χ2(2) = 4.69, p = 0.096). The model with the interaction term had a significant first/last FCNef day x Non-Consec/Low-Rew group interaction (*p* = 0.036) which indicates that the trajectory for rs-FC from day 0 to day 4 differed for the Non-Consec/Low-Rew group compared to the reference Consec/Low-Rew group (see Supplementary Table 20 and for visualisation of this see the Group Comparison inset in Figure 4).

**Figure 4.**
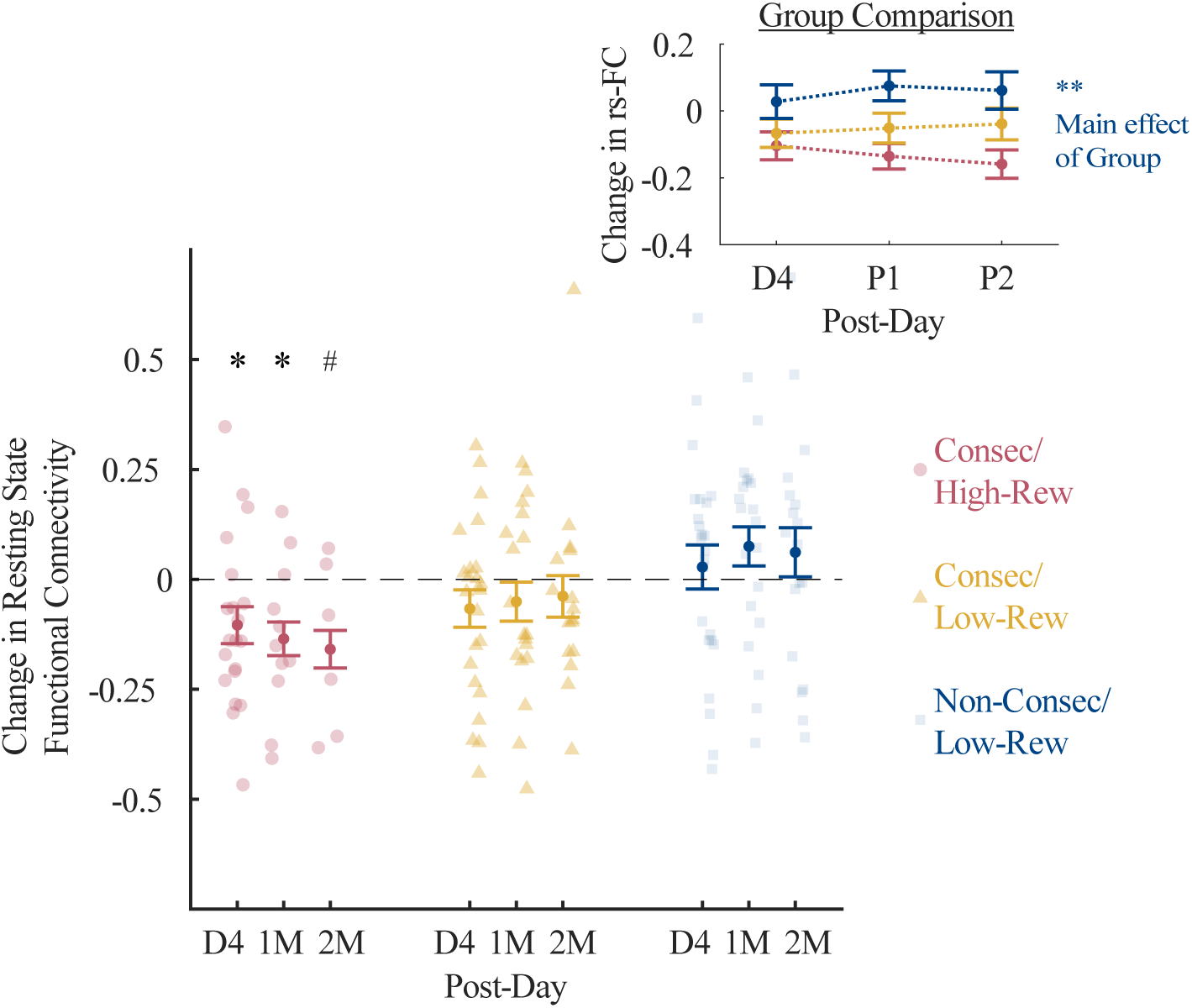
Long-term changes in rs-FC. *Investigative t-tests revealed that for the Consec/High-Rew group, overall DLPFC-PCC resting state functional connectivity (rs-FC) was significantly more negative than baseline immediately after FCNef had been completed (D4) and one-month later (M1), and that it trended toward this even two-months later (M2). Nothing of significance was found for either of the other groups, although visually, the rs-FC appears to shift in the non-targeted direction (positive) for the Non-Consec/Low-Rew group. These results should be further explored using larger sample sizes for long-term data. Note that bars for each Group represent different sample sizes because not all participants came back for long-term testing (see Table 1). The Group Comparison inset shows the mean ± standard error of the change in rs-FC, over different post-days, overlaid for the different groups. Changes = data from each post-day minus data from Day 0; D = Day, M = Month, * represents p_FDR_ < 0.05, # represents p_FDR_ = 0.051*.

##### 3.2.3.2. rs-FC: Changes in the long-term

We next examined whether any post-changes in DLPFC-PCC rs-FC differed or remained stable over time. In the best-fit model, which had no interaction term (see Table 2 and Supplementary Table 21), no significant main effect of post-day was found. This indicates that changes from baseline remained stable across post-days. A significant main effect of the Non-Consec/Low-Rew group was found (*p* = 0.007), indicating that post-changes in rs-FC differed for the Non-Consec/Low-Rew group and the reference Consec/High-Rew group. Specifically, post-changes were more negative, which was the trained direction, for the Consec/High-Rew group versus the Non-Consec/Low-Rew group (see Figure 4). Post-changes for the Consec/Low-Rew group lay between those of the other two groups (see Figure 4).

#### 3.2.4. Relationship between changes in self-report symptom scores and changes in rs-FC

Finally, in the three groups of participants, we examined how changes in participant DLPFC-PCC rs-FC from before to after FCNef related to changes in general depressive, brooding, and anxiety symptoms. Here, changes were defined as day 0 data subtracted from day 4 data. If the FCNef paradigm was successful, then we would expect changes in the DLPFC-PCC rs-FC to be positively related to changes in general depressive symptoms. This would mean that the more negative the rs-FC became, the more depressive symptoms decreased. If, as previously hypothesised [12], the targeted functional connection (FC) is specifically related to maladaptive symptoms of rumination, then we would expect changes in the DLPFC-PCC rs-FC to be positively related to changes in brooding symptoms, but not to changes in anxiety symptoms (which we used as a control). Importantly, by including Group as a factor in the models, we examined whether these effects were impacted by manipulated parameters.

##### 3.2.4.1. Relationships between symptom changes and rs-FC changes

The best-fit model to explain changes in general depressive scores (see Table 2 and Supplementary Table 22) had a significant interaction between rs-FC change and group (*p* = 0.0001). Follow-up correlations revealed a significant positive correlation between general depression change and rs-FC change for the Consec/High-Rew group (*r* = 0.66, *p_FDR_* = 0.002), a positive, but non-significant relationship for the Consec/Low-Rew group (*r* = 0.30, *p_FDR_* = 0.13), and a negative, non-significant relationship for the Non-Consec/Low-Rew group (*r* = -0.41, *p_FDR_* = 0.98).

The best-fit model to explain changes in brooding scores (see Table 2 and Supplementary Table 23) showed no significant main effects. However, hypothesis-driven follow-up correlations revealed a significant positive correlation between brooding change and rs-FC change for the Consec/High-Rew group (*r* = 0.63, *p_FDR_* = 0.003), but nothing even approaching significance for the other two groups (*r* = 0.18, *p_FDR_* = 0.31 for the Consec/Low-Rew group; *r* = 0.05, *p_FDR_* = 0.40 for the Non-Consec/Low-Rew group).

The best-fit model to explain changes in anxiety scores (see Table 2) had no significant main effects or interactions (*p*s > 0.05; see Supplementary Table 22). Follow-up correlations also showed nothing of significance (*r* = 0.05, *p_FDR_* = 0.63 for the Consec/High-Rew group; *r* = -0.08, *p_FDR_* = 0.65 for the Consec/Low-Rew group; *r* = 0.34, *p_FDR_* = 0.63 for the Non-Consec/Low-Rew group).

The relationship between symptom changes and rs-FC changes for all groups can be seen in Figure 5. Data of one participant whose general depression change was more than 3 STD lower than the group general depression change mean was excluded from the general depression change analysis (and Figure 5a). Data of another participant whose brooding change was more than 3 STD lower than the group mean was excluded from the brooding change analysis (and Figure 5b). This was because, although inclusion of these participants did not change the results, it meant the data failed to meet a crucial assumption underlying LME models (normal distribution of the residuals).

**Figure 5.**
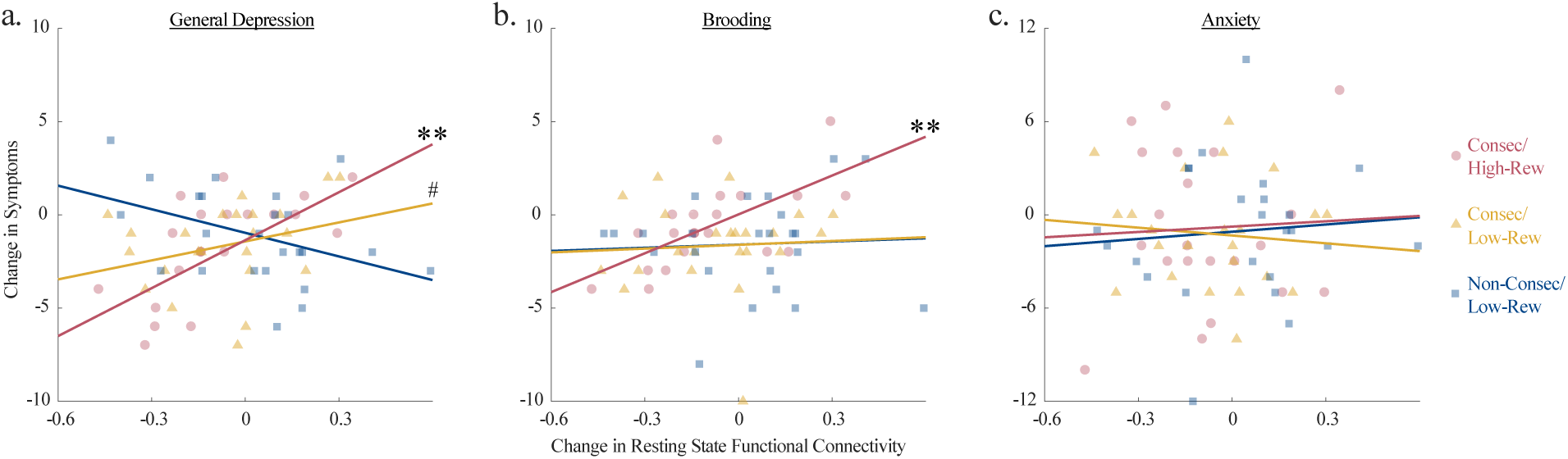
How changes in rs-FC relate to changes in symptomatology. *Changes in DLPFC-PCC resting state functional connectivity (rs-FC) are plotted against **(a)** changes in General Depression Scores, **(b)** changes in Brooding scores, and **(c)** changes in Anxiety scores. Overall, promisingly for precision medicine, the Consec/High-Rew group showed significant positive correlations between changes in rs-FC and changes in **related** (General Depression and Brooding), but not unrelated (Anxiety) symptoms. These effects were in the same direction (reaching a trend with the uncorrected p-value for the General Depression change correlation) for the Consec/Low-Rew group. On the contrary, promising results were not found for the Non-Consec/Low-Rew group, for whom the relationship between rs-FC changes and General Depression score changes were actually numerically in the opposite direction to expectation. ** represents p*_FDR_ *< 0.01, # represents p*_UNCORR_ *= 0.085*.

##### 3.2.4.2. Specificity of the targeted functional connection

We specifically hypothesised that changes in DLPFC-PCC rs-FC from before to after FCNef should relate to changes in brooding, but not anxiety symptoms. Z-tests to compare correlation coefficients [33], run with one tail due to our directional hypothesis, demonstrated a significant difference for the Consec/High-Rew group (*z* = 2.08, *p* = 0.019), but not for the other groups (*z* = 0.83, *p* = 0.21 for the Consec/Low-Rew group; *z* = 0.11, *p* = 0.46 for the Non-Consec/Low-Rew group).

## 4. Discussion

Overall, we found that the more participant DLPFC-PCC rs-FCs normalised over consecutive days of Functional Connectivity Neurofeedback (FCNef), the greater their corresponding decrease in brooding rumination, which we expected to relate to this functional connection. No such correlation was shown for changes in DLPFC-PCC rs-FC and anxiety, which is thought to relate to different underlying neural circuitry [35,36]. Importantly, here we found these results in new participants (run with consecutive days of FCNef to be consistent with our past report). Combining this newly collected data with the previously reported data provided us with sufficient statistical power for direct comparison of these correlation coefficients and a significant difference was found. When we looked at data from three groups of participants (see Table 3), we found the most promising results for the group run with a higher-reward schedule to reinforce the targeted shift in functional connectivity, and an experimental schedule with consecutive days. Weaker results were found for the group of participants that completed FCNef over consecutive days, but who were reinforced with a lower reward schedule. FCNef did not seem to have any effect on the group of participants who attended over non-consecutive days and that were reinforced with a lower reward schedule.

**Table 3:**
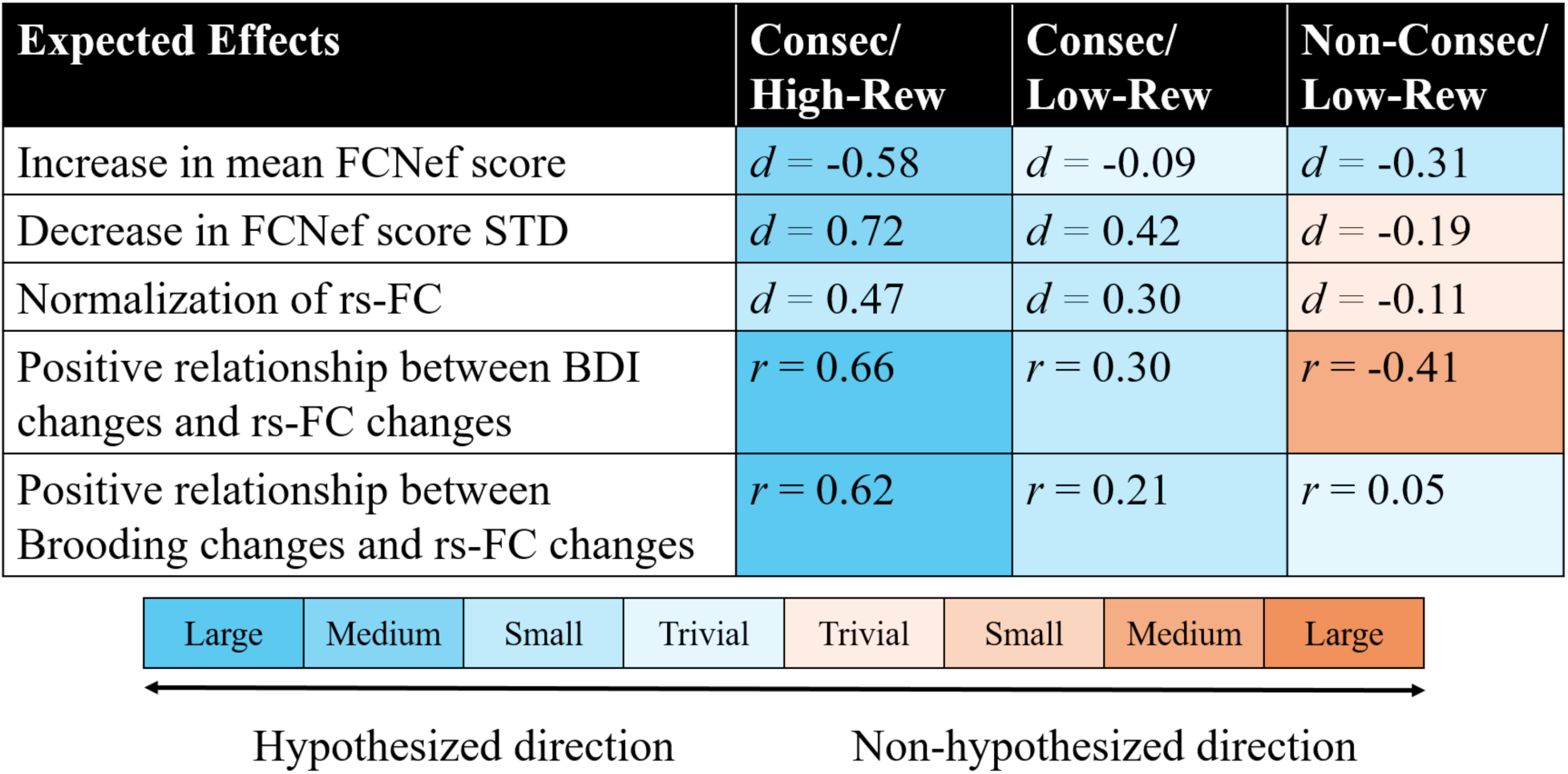
Effect sizes summarized. “Expected effects” are effects that we expect to see if FCNef training is successful. Comparisons for “Increase in mean FCNef scores” and “Decrease in FCNef score STD” were t-tests comparing data from the first and the last day of FCNef (Days 1 and 4). Comparisons for “Normalization of rs-FC” were t-tests comparing rs-FC from before to after FCNef (Days 0 and 4). The two “Positive relationship…” comparisons were correlations calculated between changes in rs-FC and changes in symptoms, where “changes” were defined as Day 4 - Day 0 data. Effect sizes for each comparison are shown in the relevant cell. These are shown here as Cohen’s d (d) for t-tests and Pearson’s correlation coefficient (r) for correlations. Following convention, these can be described on a scale ranging from trivial (d < |0.2| or r < |0.1|), small (0.2 ≤ |d| < 0.5 or |0.1| ≤ r < |0.3|), medium (|0.5| ≤ d < |0.8| or |0.3| ≤ r < |0.5|), to large (d ≥ |0.8| or r ≥ |0.5|). Effects in the hypothesized direction are shown in shades of blue and effects in the non-hypothesized direction are shown in shades of orange. The larger the effect size, the stronger the shade of the colour in the relevant cell.

### 4.1. FCNef for precision medicine

The current results strengthen our previous finding that when FCNef is run over consecutive days, normalisation of a target functional connection relates to a specific reduction in only related symptoms (see Figure 2). This replication of previous results with an independent sample of newly collected data shows their robustness and indicates that previous results were unlikely to have been spurious [12]. Furthermore, the finding that changes in the targeted rs-FC correlated significantly more with changes in related than unrelated symptoms highlights the precision of the FCNef technique. This brings us one step closer to a future in which patients may one day enter a clinic, have their brains scanned, and then have targeted treatment to normalise specific neural aberrations related to their own subset of symptoms.

### 4.2. The DLPFC-PCC functional connection and rumination

Not only do our results have implications for precision medicine in general, but also specifically for understanding and treating brooding rumination symptoms. We previously hypothesised, based on both data- and hypothesis-driven evidence from past studies, that the DLPFC-PCC functional connection is likely to relate to brooding rumination [11]. Our initial study found preliminary support consistent with this proposal. Specifically, we found that changes in connectivity between these regions relate to changes in brooding rumination [12]. There has since been at least one other report that also showed increased connectivity between the PCC and PFC related to ruminative symptoms [37]. We here find even stronger support that brooding rumination relates to the DLPFC-PCC functional connection, by showing that the correlation between changes in these is significantly stronger than the correlation between changes in the same functional connection and a type of symptom thought to be unrelated (anxiety). Evidence from previous studies using repetitive transcranial magnetic stimulation to target the DLPFC [38,39] and using real-time neurofeedback to target the PCC [40], have also shown promising results for amelioration of depressive and brooding rumination symptoms. This could be because targeting these regions affects the functional connection between them, which if true, could mean that targeting the functional connection itself more directly (as we did here) could be of even further advantage.

### 4.3. Parameter testing

The current report extends our previous report by clarifying some of the parameters under which FCNef for depression can best be achieved. These results should help guide the design of future neurofeedback and other BMI studies. When selecting parameters for neurofeedback, past studies have tended to follow convention or have gone with what seemed best in terms of cost/benefit trade-offs and/or in terms of making things easy for participants. Often, this is the only feasible way to design a BMI study because the cost of testing all possible parameters is enormous. However, our current results show that certain parameters can make a difference in BMI effectiveness. This means that without knowing the optimal parameters for a given BMI design, researchers may be finding null results simply because they are not running their designs in the most effective way. There is no simple solution to this problem, especially because optimal parameters may differ for different populations, target neural activities, experimental goals, etc. Nonetheless, the current results provide initial evidence that can be used to help future designs. Below, we discuss specific results for these parameter analyses, as well as their implications.

#### Reward schedule

Participants run with the high-reward schedule had better FCNef success than those run with the low-reward schedule (see Table 3 and Figures 3, 4, and 5). These results support the proposal that external reward might work as reinforcement that is additional to that provided by feedback scores during neurofeedback [41]. Based on these results, we recommend using liberal external reward in future neurofeedback studies. Furthermore, because BMIs generally do not use external rewards for reinforcement, this result might be worthy of consideration beyond the realm of neurofeedback.

Of course, disturbances to reward circuitry and disturbances in reward processing (usually reductions) are commonly reported in depressive and other psychiatric disorders [42–49]. This means that the effect of external reward on reinforcement of the target neural activity might be diminished when neurofeedback is conducted in patients with such disorders. Here, our results with subclinical patients did not corroborate this, but it remains worthy of further investigation in clinically depressed patients.

#### Experimental schedule

FCNef appears to be more effective when participants come for consecutive, as opposed to non-consecutive, days of FCNef. All expected effects were strongest in the consecutive condition with high reward, in the same direction (albeit with less strength) in the consecutive condition with low reward, and weak or in the opposite direction for the non-consecutive condition (see Table 3). Of course, the non-consecutive condition that we tested was with low reward and it would have been nice to fully balance this by also testing the non-consecutive condition with high. reward. While these results are therefore not fully conclusive, they do not appear promising for using non-consecutive days of FCNef. They indicate that, although possibly more tiring for participants, consecutive days of FCNef may be necessary to achieve positive outcomes. It is possible that consecutive days of reinforcement are needed to drive learning effectively and/or that more non-controllable confounding personal circumstances can occur between non-consecutive days of training (an idea that researchers designing future BMIs and clinical treatments ought to consider).

Another point to consider is the possibility that neural plasticity related to learning might cause dynamic rs-FC fluctuations in strength and direction that occur before settling into a new pattern (similar to the rebound effect first documented by Kluetsch et al., 2014 [34]). If so, then our analyses may not have fairly tested consecutive versus non-consecutive conditions. Learning could begin from Day 1; but, post-FCNef measurements (from Day 4, and 1- and 2-months later) differed in the number of days after Day 1 for consecutive/non-consecutive conditions (and even for different participants in the non-consecutive condition). This may mean that measurements were taken at different points during ongoing dynamic rs-FC fluctuations for these different conditions. If so then comparisons between these conditions may actually have compared different snapshots of learning effects. Other researchers using neurofeedback over non-consecutive days should also consider this possibility.

### 4.4. Limitations of the current design

One limitation of our study is that it involved participants who had only subclinical levels of depression. Nonetheless, preliminary studies using this FCNef technique with patients diagnosed with MDD have shown promise [19,21]. Results with clinical patients may be improved if the right parameters are employed. The most effective parameter that we found was high(er) monetary reward. We used money because we wanted to test the effects of reward schedule with a type of reward that is well known to strongly activate the human reward system. However, now that proof-of-concept has been provided, this idea should be further tested more creatively with other types of reward that might be more appropriate for a clinical setting. For example, revealing consecutive puzzle pieces for each successful trial of neuromodulation (see Ramot et al., 2017 [16]). A second limitation of our study is the absence of a control group or within-subject control condition. Therefore, it is possible that our target rs-FC changed and that symptoms improved for reasons such as the placebo or Hawthorne effects. However, only changes in relevant (depressive and brooding rumination, but not anxiety) symptoms changed parallel to the targeted FC, which would be unlikely to occur merely from such nonspecific effects. A third limitation of our study is the incomplete factorial design due to practical constraints, as discussed in the Experimental Conditions section. While this limits our ability to examine potential interaction effects between the experimental schedule and reward schedule parameters, the current design still allows us to investigate main effects of these parameters separately, which was our main aim. Future research should include all factorial combinations to provide a comprehensive understanding of parameter interactions.

### 4.5. Future directions

Although different symptoms are likely to arise from aberrations in wider brain networks involving multiple FCs, our current FCNef approach can directly target only one FC. In that sense, connectome-based FCNef [50] or neurofeedback targeting an estimation of the dynamic weighted linear sum of FCs might be more effective. Some promise for such types of neurofeedback has been found, including when targeting an estimation of the dynamic weighted linear sum of FCs from the greater biomarker from which we identified the DLPFC-PCC FC [T. Ogawa, personal communication, 27^th^ January, 2025; 51]. However, the authors have also reported increased difficulty for participants with regard to the credit assignment (they report that it is difficult to target multiple functional connections and to know what actually worked) and overall experimental interpretability [T. Ogawa, personal communication, 27^th^ January, 2025]. Furthermore, FCNef itself affects broader brain networks than just the targeted functional connection anyway [15,16], so it remains possible that our simple FCNef approach might be best at ameliorating symptoms without the need for added complexity and burden for patients. Future studies should attempt to directly compare effectiveness of these types of neurofeedback.

Our current results are promising for precision medicine, but they were shown with functional connectivity in fMRI, which can be costly and impractical (but not impossible) for real clinical treatment. In the future, we expect FCNef to evolve further so that it may be conducted using electroencephalogram (EEG) signatures (see Keynan et al., 2019 [52]) of target FCs or so that it may be conducted using EEG signatures of weighted linear sums of multiple FCs. If successful, then this would allow neurofeedback targeting functional connections to eventually be conducted with portable EEG headsets, possibly even away from the clinic in the privacy of the patient’s own home (for more detailed discussion see Taylor et al., 2021 [53]).

### 4.6. Conclusion

Overall, we have extended previous results to show that normalisation of the targeted neural network (DLPFC-PCC) correlated significantly more with reductions in symptoms thought to relate to this neural circuitry (brooding rumination) than to changes in symptoms thought to relate to different neural circuitry (anxiety). This highlights the precision of the FCNef technique and brings us one step closer to a future where psychiatric treatment might be tailored to the individual patient. Here, we additionally extended our previous work by investigating parameters under which our FCNef for depression paradigm is most effective. We found that FCNef effectiveness changes depending on those parameters with which it was run. Specifics and implications of some parameter-related results may be relevant beyond neurofeedback to BMIs in general. Furthermore, some of our results highlight benefits of testing conventional parameters. Overall, these results should be informative for design of future BMI testing and for inspiring new interpretations of existing data. For example, previously found null results should be considered in the context of the parameters under which the BMI was run. More broadly, by documenting how parameter optimisation can increase beneficial outcomes and reduce patient burden, we hope to inspire more of this in the future, with the ultimate goal of bringing optimised BMIs to the medical clinic.

## Supporting information

MS Word document of manuscript with no figures or tables

## Data Availability

All data produced in the present study are available upon reasonable request to the authors

## Acknowledgements

We thank Kaori Nakamura for help in scheduling and conducting the experiments, Rumi Yorizawa for help in screening participants, and Toshinori Yoshioka for continued support with regard to experimental scripts.

## Statement of ethics

Participants all provided written informed consent on each day of screening and experimentation prior to commencement. This research was approved by the Ethics Committee of the Review Board of Advanced Telecommunications Research Institute International, Japan (Ethics No. 132, 172) and by the Kyoto University Certified Review Board (YC0849) and the Committee on Medical Ethics of Kyoto University (C0849). All experiments were performed in accordance with the guidelines and regulations of these Ethics Committees. This research was conducted in association with research registered with the Japan Registry of Clinical Trials (jRCTs052180169) and the University Hospital Medical Information Network Clinical Trials Registry (UMIN000015249).

## Author Contributions

JET, TM, TY and MK designed experiments; JET, TO, MM and TM acquired data; JET analysed data; JET prepared the original draft; TO, MM, MT, TY, TK, YK, YY, JM, TM, MK, and AC reviewed and edited the manuscript. All authors gave final approval for submission and agreed to take responsibility for the manuscript.

## Data accessibility

Data and code supporting this study’s findings will be publicly available on our GitHub at publication.

