## Supplementary Files for "Paving the way for precision treatment of psychiatric symptoms with functional connectivity neurofeedback"

### Supplementary Materials

#### Methods

##### Experimental Procedure

Functional localiser day

⇒ *outside the scanner*

1. Questionnaire Battery: Participants completed a battery of questionnaires, including the Beck Depression Inventory-II (BDI)[1], Rumination Response Scale (RRS)[2,3], and Trait Anxiety Scale (STAI-Y2)[4].

⇒ *inside the scanner*

2. Resting-state fMRI: Participants were instructed to focus on a central fixation point on a monitor and to relax. Scanning took 10 min.
3. T1-weighted structural MRI: Participants were again instructed to focus on a central fixation point on a monitor and to relax. Scanning took 9 min and 50 sec.
4. N-back task. Over 24 blocks (split into three sessions), participants saw a series of numbers with intermittent rest periods. Depending on the block, participants were instructed to press a button any each time they saw a number ("0-back") or each time the presented number was the same as the number presented one, two, or three numbers earlier ("1-back", "2-back", and "3-back", respectively). Recruitment of the Executive Control Network is expected to increase with task difficulty (i.e. 3-back>2-back>1-back>0-back) (Thompson et al., 2016). Conversely, recruitment of the Default Mode Network is expected to be highest during rest periods (Raichle et al., 2001). We thus used this task to localise peak DLPFC/mFG activity for each participant under conditions of expected high Executive Control Network recruitment and to localise peak PCC/precuneus activity for each participant under conditions of expected high Default Mode Network recruitment. These localisations were then used to build individualised regions of interest for each participant (specific analysis details in Taylor et al., 2022[5]). The total scanning time for this task was 18 min and 39 sec.

#### Day 0

⇒ *inside the scanner*

1. SHAM task (Figure. 1, see Taylor et al. 2022[5] for more details): participants performed the FCNef task with random feedback (rather than feedback based on their neural activity). The purpose was to determine a baseline level of functional connectivity for each participant between the two target ROIs (DLPFC-PCC) with which we could compare their functional connectivity from the FCNef task. Importantly, participants did not know that the feedback was random. They had been instructed that the feedback depended on their neural activity from during induction period of the task.
2. Resting-state fMRI

⇒ *outside the scanner.*

3. Questionnaire Battery: Same as above.

#### FCNef days 1-4

⇒ *inside the scanner*

1. FCNef task (Figure 1, see Taylor et al. 2022[5] for more details): On Day 1, participants received an apology and an explanation that the feedback on the previous day had been random, but that it would be real henceforth. On all days of the FCNef task (Days 1-4), participants were instructed that the feedback would be based on their neural activity from the induction period of the task, and importantly, it really was. Participants were given no clue as to what specific neural activity this feedback was based on, but in reality it was based on functional connectivity between the targeted regions of interest. Specific details of the feedback calculation are detailed below in the section entitled 'FCNef and SHAM score calculation.' This feedback corresponded to a real monetary reward bonus, which differed between conditions and is described in detail in the section in the main text entitled, "Differences in Experimental Procedure".
2. Resting-state fMRI: Same as above.

⇒ *participants excited the scanner.*

3. Questionnaire Battery (only on FCNef Day 4): Same as above.

#### Follow-up (Post) testing (one and two months after FCNef)

⇒ *participants entered the scanner.*

1. Resting-state fMRI: same as above.

*⇒ participants excited the scanner.*

2. Questionnaire Battery: same as above.

### Participants

#### Participant screening and recruitment

Participants were first screened and then, when appropriate, recruited for the main experiment. Recruitment for screening was done in various ways, such as via leaflets sent via mail, poster advertisements in major local railway stations and universities, online job-seeking websites, and by word-of-mouth. Our only limitations for screening were that participants must be 20-40 years old and must speak Japanese. Screening took place over two days, with at least one week between. On each screening day, participants completed the Beck Depression Inventory-II (BDI) questionnaire[1]. Participants were deemed as potentially able to participate in the main experiment if (a) they had a mean BDI score $\geq$ 8, averaged over the two days of screening, and (b) if they indicated no intention to commit suicide. Participants who met these criteria next met with a medical doctor or clinical psychologist, who determined whether they currently had a diagnosis of or were receiving treatment for a psychiatric illness. Such participants were excluded due to ethical considerations, as we did not yet know the full extent of short and long-term effects of FCNef manipulation under the range of conditions used in this study. Otherwise, participants were invited back for the main experiment.

#### Participant demographic information

Here we report data collected from 68 participants, 19 of whom have had their data reported in a previous study. Including data from those 19 participants in the present study allowed us to better explore contrasts between our three parameters of interest. These 68 participants all had normal or corrected-to-normal vision. They had a mean age of 25.94 (std = 6.67), and 32 identified as female. There were no significant differences in ages between groups (mean = 27.38, std = 7.14, for Consec/High-Rew; mean = 25.09, std = 6.58, for Consec/Low-Rew; mean = 25.50, std = 6.42, for < 0.05 Non-Consec/Low-Rew; $ps_{FDR} > 0.05$). There were no significant differences between sex for the Consec/High-Rew group (9/21 were female) and the other groups ($ps_{FDR} > 0.05$). There was a significant difference in sex for the other two groups ($t(45) = 2.60$, $p_{FDR} < 0.05$; 15/23 were female for Consec/Low-Rew, 7/24 were female for Non-Consec/Low-Rew).

Because we found significant gender differences between the Consec/Low-Rew and Non-Consec/Low-Rew groups, we re-ran best-fit models from all analyses (see Table 2 of the main text) with

Sex as an additional IV to see if this could explain any of our findings. In the case that the best-fit model was the multiplicative model, interactions with Sex were also included as IVs. Likelihood ratio tests showed that adding this/these IV/IVs significantly improved only one of the best-fit models. This means that with one exception, sex was not a meaningful predictor of results. This one exception was the model to predict General Depression (BDI) scores in the short-term (General Depression ~ first/last FCNef Day + Group + **Sex** + (1|Subject)' was significantly better than General Depression ~ first/last FCNef Day + Group + (1|Subject)'; $\chi^2(1) = 7.57$, $p = 0.006$). Follow-up tests showed that General Depression scores were higher for females (m = 13.56, std = 6.61, on Day 0; m = 11.38, std = 6.07, on Day 4) than males (m = 9.69, std = 5.85, on Day 0; m = 8.64, std = 5.58, on Day 4) on the first and last days of FCNef. However, this additive model did not include an interaction between Group and Sex. If such an interaction was significant, then the multiplicative model would have been statistically better than this model, and so we conclude that **differences in sex did not meaningfully cause differences in General Depression scores** on Day 0 and Day 4 **between groups.**

#### Participant payment

For each questionnaire session (including screening sessions), participants were paid ¥3000. They were paid a base reward of ¥8000 for each MRI session. In SHAM and FCNef sessions, they also received a reward bonus, as described in the main text.

**Supplementary Table 1.**

*This table is the same as Table 1 from the main text, but with sample sizes for each group categorised depending on whether participants were run with 20 or 40s induction time-windows.*

| Group | Induction Time-Window | Main Experiment Sample Size | 1-Month Follow-up Sample Size | 2-Month Follow-up Sample Size |
|---|---|---|---|---|
| Consec/High-Rew | 40s | 9 | N/A | N/A |
| | 20s | 12 | 11 | 9 |
| Consec/Low-Rew | 40s | 10 | 9 | 8 |
| | 20s | 13 | 11 | 9 |
| Non-Consec/Low-Rew | 40s | 12 | 11 | 9 |
| | 20s | 12 | 11 | 11 |
| Total | | 68 | 53 | 46 |

### Materials

The protocol was the same as reported in Taylor et al. (2022)[5]. We quote the relevant part of that publication: "*Visual stimulus presentation was controlled using MATLAB 7.5.0.342 (2007b; The MathWorks Inc.). The visual stimuli were projected to an opaque screen inside the scanner via a projector (DLA-X7-B, JVC; frame rate = 60 Hz) and a mirror system. Participants responded to the stimuli using MRI-compatible response pads (HHSC-2 × 2, Current Designs, Inc., PA, USA).*"

#### Self-report symptom questionnaires

##### Beck Depression Inventory-II (BDI)

The BDI comprises 21 items rated on a four-point scale from 0 to 3. Higher BDI scores indicate greater severity of general depressive symptoms. Reliability and validity have been confirmed[1].

##### Rumination Response Scale (RRS)

The RRS comprises 22 items rated on a four-point scale from 1 (never) to 4 (almost always). It measures rumination symptoms; i.e., symptoms related to repeated self-reflection about negative emotions. For analysis, we used only the brooding subscale of this questionnaire. This consists of 5 items related to the maladaptive form of rumination ("moody pondering") when this is unconfounded by general depressive symptoms[2]. Reliability and validity have been confirmed.[2,3]

##### State-Trait Anxiety Inventory (STAI)

The STAI comprises 40 items rated on a four-point scale from 1 (not at all) to 4 (very much), which measures general anxiety symptoms. For analysis, we used only the trait anxiety subscale of this questionnaire (STAI-Y2), which comprises 20 of these items. Reliability and validity have been confirmed.[4]

#### Imaging data acquisition

A 3T MR scanner with a 32-channel head coil, located at the ATR Brain Activity Imaging Centre, was used for data acquisition (Siemens MAGNETOM Verio, Siemens, Erlangen, Germany). Anatomical images were acquired using a T1-weighted MP-RAGE protocol (slice number, 240; matrix size, 256 * 256; FOV, 256 mm; voxel size, 1.0 * 1.0 * 1.0 mm (no slice gap); TR, 2300 ms; TE, 2.98 ms; flip angle, 9°). T2*-weighted images reflecting blood oxygen level-dependent (BOLD) signals were acquired in all experimental and resting-state sessions using gradient-echo echo-planar imaging (EPI) (slice number, 60; matrix size, 100 * 100; FOV, 200 mm; voxel size, 2.0 * 2.0 * 2.0 mm (0.5mm slice gap); TR, 1000 ms; TE, 28 ms; flip angle, 65°). Multiband sequence, with an acceleration factor of 6, allowed for faster slice acquisition [6–8]. Each 'functional localiser task' session took 590s and consisted of 590 volumes. Each SHAM FCNef and FCNef session took 512s and consisted of 512 volumes. Each resting-state session took 600s and consisted of 600 volumes. The first ten volumes taken in each session of all experimental and resting-state sessions were discarded to ensure steady-state magnetisation.

#### FCNef and SHAM score calculation

All participants received a baseline reward bonus of ¥500 on each day for the SHAM and FCNef tasks. On each day of FCNef (but not on the SHAM day), they could receive an additional reward bonus. This depended on their average FCNef scores from that day. FCNef scores were calculated based on the targeted functional connection from the induction period of each trial. If this was the same as participant mean baseline functional connectivity (that from the SHAM task on Day 0) then the FCNef score on that trial would be 50. If this was more anti-correlated than participant mean baseline functional connectivity, then the FCNef score on that trial would be higher than 50, up to a score of 100 (which was calculated as the mean baseline FC - the std baseline FC). This is because we wished to reinforce more negative connectivity. On the contrary, if the FC was more positively correlated than participant mean baseline functional connectivity, then the FCNef score on that trial would be lower than 50, down to a score of 0 (which was calculated as the mean baseline FC + the std of baseline FC). All task scores were presented as a green feedback circle on screen, the maximum circumference of which corresponded to a score of 100 and the minimum circumference of which corresponded to a score of 0 (Figure 2). Note that SHAM scores (scores from the SHAM task on Day 0) were random (see [5]) and were not used to calculate reward.

**Supplementary Table 2.**

*An example of reward bonus payments for different average daily FCNef scores, depending on the (low/high) reward schedule condition, is displayed.*

| FCNef Score | Low Reward Bonus | High Reward Bonus |
|---|---|---|
| 50 | ¥500 | ¥500 |
| 60 | ¥500 | ¥1,000 |
| 70 | ¥500 | ¥1,500 |
| 80 | ¥1,000 | ¥2,000 |
| 90 | ¥2,000 | ¥2,500 |
| 100 | ¥3,000 | ¥3,000 |

### Supplementary Results

Due to space restrictions of tables presented hereafter, groups of participants will be referred to as C/HR (the Consec/High-Rew group from the main text), C/LR (the Consec/Low-Rew group from the main text), and NC/LR (the Non-Consec/Low-Rew group from the main text).

#### Pilot study/analysis investigating induction time-window

We re-analysed FCNef data from the 40s time-window of the induction period of 15 participants who were run in preliminary versions of our FCNef for depression paradigm (with a consecutive-day experimental schedule and high reward). As in all versions of this design to date, participants were reinforced during FCNef when they made their DLPFC-PCC FC more negative than their baseline FC (which was calculated from SHAM data). We wished to examine how “effectively” participants were using the 40s time-window and so we analysed different “time intervals” from within it.

Here we show results for the following time-intervals of the induction period: 0-10s, 10-20s, 20-30s, and 20-40s. The DLPF-PCC FC was calculated (with the same method as during online FCNef, described in Taylor et al., 2022[5]) for each of these time-intervals. This was put into LME models as the dependent variable. Independent variables of “TI” (Time Interval: 0-10s, 10-20s, 20-30s, and 20-40) and “Day” (Day 0, 1, 2, 3, or 4) were included in the model. A model which included additive effects of these variables (‘FC~Day+TI’; AIC=-229.10) was not significantly improved by including the interaction between them (‘FC~Day*TI’; AIC=-229.87) ($\chi2(1) = 2.77$, $p = 0.100$). It was improved by adding a random intercept for experimental subject (‘FC~Day+TI+(1|Subject)'; AIC=-266.01) ($\chi^2(1) = 38.91$, $p < 0.001$).

The best-fit model had a significant main effect of Day and a significant main effect of TI (see Supplementary Table 3, below). These results reflect the fact that overall average DLPF-PCC FCs changed over days of FCNef and that they were different for different TIs. As can be seen in Supplementary Figure 1 (below), consistent with successful FCNef, participant DLPF-PCC FCs became more negative over FCNef days. Interestingly, as is most apparent in the results for FCNef Days 3 and 4 (reflecting an effect of learning), these DLPF-PCC FCs were more negative in 0-10s and 10-20s TIs than they were in 20-30s and 30-40s TIs. Because the goal was to make DLPF-PCC FCs more negative, we concluded that participants learned most effectively to do this in the first 20s of the 40s induction time-window. When we compared the DLPF-PCC FCs on each day of FCNef as analysed from a full 20s time-window (from 0-20s) compared to the full 40s time-window (from 0-40s) we found no significant differences (uncorrected *p*s >0.05; bear in mind that data from the 0-20s time-window were also included in the 0-40s time-window). Overall, therefore, it seems that participants most effectively used the first 20s of the 40s time-window and that DLPF-PCC FCs did not change depending on whether a 20s or 40s time-window was used for analysis. Results of this analysis were our basis for testing 20s versus 40s induction time-windows experimentally (see Supplementary Table 1 and the results reported below).

**Supplementary Table 3. Best-fit LME to predict average online DLPFC-PCC FCNef from pilot data.**

*Model: FC~ TI + Day +(1|Subject)*

*Main factor variables: FC= linear, TI = linear, Day = linear*

| Variables | Estimate | Lower | Upper | SE | *t* Stat | DF | *p* Value |
|---|---|---|---|---|---|---|---|
| (Intercept) | -0.18 | -0.24 | -0.12 | 0.03 | -5.95 | 297 | < 0.001 |
| TI | 0.00 | 0.00 | 0.00 | 0.00 | 3.05 | 297 | 0.003 |
| Day | -0.05 | -0.06 | -0.04 | 0.01 | -8.23 | 297 | < 0.001 |

**Supplementary Figure 1. DLPFC-PCC FC from different "time intervals" (TIs) of a 40s induction time-window.**

*Overall, DLPF-PCC FCs become more negative over FCNef training days. They were particularly more negative in the 0-10s and 10-20s TIs on Days 3 and 4. D = Day. FC = functional connectivity between the DLPFC and PCC.*

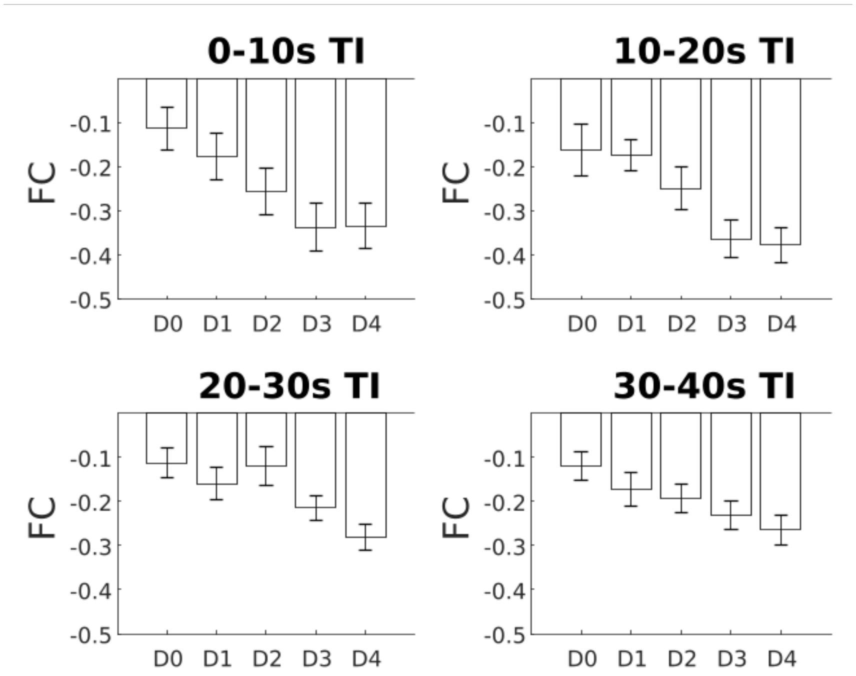


### No influence of time-window on main results reported for the total dataset of the current report

We ran LME models to examine how changes in participant DLPFC-PCC rs-FC from pre- to post-FCNef related to changes in symptoms. These models were run with and without the interaction term between independent variables and we tested these against one another to see which best fit the data.

No interaction term: *Symptom Change~rs-FC Change+Group+Time-Window+(1|Subject)'*

With interaction term: *Symptom Change~rs-FC Change*Group*Time-Window+(1|Subject)'*

The best-fit model to predict changes in general depressive symptoms was the model that included the interaction term (see Supplementary Table 4). Similar to the model reported in the main text, this model had a significant interaction between rs-FC change and group ($p$ = 0.0001). Note that there were no main effects or interactions with Time-Window ($p$s > 0.05).

The best-fit models to predict changes in brooding and anxiety symptoms were models that did not include the interaction term (see Supplementary Tables 5 and 6). Like those models reported in the main text, these models had no significant main effects ($p$s > 0.05). Note here that this also means there was no significant main effect of Time-Window in either case.

Because these best-fit models showed no important contribution of induction time-window to these main results, we did not include induction time-window in models reported in the main text.

**Supplementary Table 4. Best-fit LME to predict General Depression Change (Day 4 - Day 0) using rs-FC Change (Day 4 - Day 0), Group, and Time-Window.**

*Model: General Depression Change ~ rs-FC Change x Group x Time-Window + (1|Subject)*

*Main factor variables: General Depression Change = linear, rs-FC Change = linear, Group = categorical, Time-Window = linear*

| Name | Estimate | Lower | Upper | SE | *t* Stat | DF | *p* Value |
|---|---|---|---|---|---|---|---|
| (Intercept) | -1.14 | -3.66 | 1.38 | 1.26 | -0.09 | 58 | 0.368 |
| Time-Window | -0.77 | -4.31 | 2.78 | 1.77 | -0.43 | 58 | 0.667 |
| rs-FC change | 23.33 | 12.19 | 34.47 | 5.57 | 4.19 | 58 | < 0.001 |
| Group | -0.10 | -1.18 | 0.99 | 0.54 | -0.18 | 58 | 0.860 |
| Time-Window*rs-FC Change | -12.36 | -28.56 | 3.38 | 8.09 | -1.53 | 58 | 0.132 |
| Time-Window*Group | 0.51 | -1.02 | 2.04 | 0.76 | 0.67 | 58 | 0.505 |
| rs-FC Change*Group | -9.74 | -14.53 | -4.94 | 2.39 | -4.07 | 58 | 0.0001 |
| Time-Window*rs-FC Change*Group | 5.37 | -1.41 | 12.15 | 3.39 | 1.59 | 58 | 0.118 |

**Supplementary Table 5. Best-fit LME to predict Brooding Change (Day 4 - Day 0) using rs-FC Change (Day 4 - Day 0), Group, and Time-Window.**

*Model: Brooding Change ~ rs-FC Change + Group + Time-Window + (1|Subject)*

*Main factor variables: Brooding Change = linear, rs-FC Change = linear, Group = categorical, Time-Window = linear*

| Name | Estimate | Lower | Upper | SE | *t* Stat | DF | *p* Value |
|---|---|---|---|---|---|---|---|
| (Intercept) | -0.05 | -2.25 | 1.33 | 0.89 | -0.52 | 62 | 0.608 |
| Time-Window | 0.09 | -1.05 | 1.23 | 0.57 | 0.16 | 62 | 0.873 |
| rs-FC change | 1.76 | -0.89 | 4.41 | 1.33 | 1.33 | 62 | 0.189 |
| Group | -0.44 | -1.17 | 0.29 | 0.36 | -1.22 | 62 | 0.228 |

**Supplementary Table 6. Best-fit LME to predict Anxiety Change (Day 4 - Day 0) using rs-FC Change (Day 4 - Day 0), Group, and Time-Window.**

*Model: Anxiety Change ~ rs-FC Change + Group + Time-Window + (1|Subject)*

*Main factor variables: Anxiety Change = linear, rs-FC Change = linear, Group = categorical, Time-Window = linear*

| Name | Estimate | Lower | Upper | SE | *t* Stat | DF | *p* Value |
|---|---|---|---|---|---|---|---|
| (Intercept) | -0.18 | -3.78 | 3.41 | 1.80 | -0.10 | 63 | 0.919 |
| Time-Window | -1.48 | -3.78 | 0.27 | 1.15 | -1.28 | 63 | 0.205 |
| rs-FC change | 1.28 | -4.10 | 6.66 | 2.69 | 0.47 | 63 | 0.637 |
| Group | -0.28 | -1.75 | 1.18 | 0.73 | -0.39 | 63 | 0.699 |

**Supplementary Table 7. Comparisons of baseline measurements (taken on Day 0) for different groups of participants.**

| Group Comparison | rs-FC | | BDI | | Brooding | | Anxiety | |
|---|---|---|---|---|---|---|---|---|
| | *t* Stat | *p* FDR | *t* Stat | *p* FDR | *t* Stat | *p* FDR | *t* Stat | *p* FDR |
| C/HR vs. C/LR | 1.20 | 0.298 | 0.86 | 0.624 | -1.1 | 0.441 | 1.22 | 0.230 |
| C/HR vs. NC/LR | 2.01 | 0.153 | 0.27 | 0.792 | 0.15 | 0.880 | -1.87 | 0.102 |
| C/LR vs. NC/LR | 1.05 | 0.298 | -0.8 | 0.624 | 1.47 | 0.441 | -3.75 | 0.002 |

**Supplementary Table 8. Best-fit LME to predict Mean FCNef scores.**

*Model: Mean Score ~ FCNef Day + Group + (1|Subject)*

*Main factor variables: Mean Score = linear, FCNef Day = linear, Group = categorical*

| Variables | Estimate | Lower | Upper | SE | *t* Stat | DF | *p* Value |
|---|---|---|---|---|---|---|---|
| (Intercept) | 52.79 | 46.52 | 59.05 | 3.18 | 16.59 | 259 | < 0.001 |
| FCNef Day | 1.91 | 0.79 | 3.04 | 0.57 | 3.35 | 259 | 0.0009 |
| C/LR | -5.79 | -13.53 | 1.95 | 3.93 | -1.47 | 259 | 0.142 |
| NC/LR | -7.97 | -15.80 | -0.14 | 3.98 | -2.01 | 259 | 0.046 |

**Supplementary Table 9. Best-fit LME to predict variance (standard deviations, STD) in FCNef scores.**

*Model: STD Score ~ FCNef Day * Group + (1|Subject)*

*Main factor variables: Mean Score = linear, FCNef Day = linear, Group = categorical*

| Variables | Estimate | Lower | Upper | SE | *t* Stat | DF | *p* Value |
|---|---|---|---|---|---|---|---|
| (Intercept) | 39.51 | 36.95 | 42.07 | 1.30 | 30.37 | 257 | < 0.001 |
| FCNef Day | -1.73 | -2.54 | -0.91 | 0.42 | -4.16 | 257 | < 0.001 |
| C/LR | -0.62 | -4.16 | 2.93 | 1.80 | -0.34 | 257 | 0.732 |
| NC/LR | -2.46 | -6.07 | 1.14 | 1.83 | -1.34 | 257 | 0.180 |
| FCNef Day:C/LR | 0.93 | -0.20 | 2.06 | 0.57 | 1.62 | 257 | 0.107 |
| FCNef Day:NC/LR | 1.94 | 0.79 | 3.09 | 0.58 | 3.33 | 257 | 0.001 |

**Supplementary Table 10: Average ± standard deviation of questionnaire scores on different days of measurement for the three groups.**

| Group | Day | General Depressive Score | Brooding Score | Anxiety Score |
|---|---|---|---|---|
| C/HR | Day 0 | 12.33 ± 1.83 | 10.52 ± 0.86 | 44.29 ± 2.53 |
| | FCNef Day 4 | 10.23 ± 1.44 | 9.95 ± 0.89 | 43.38 ±2.91 |
| | Post 1-month | 10.73 ± 1.80 | 7.55 ± 0.43 | 36.45 ± 2.11 |
| | Post 2-months | 11.11 ± 1.89 | 7.78 ± 0.39 | 40.33 ± 2.25 |
| C/LR | Day 0 | 10.48 ± 1.22 | 11.78 ± 0.81 | 40.69 ± 1.62 |
| | FCNef Day 4 | 8.83 ± 1.15 | 10.13 ± 0.81 | 37.56 ± 1.53 |
| | Post 1-month | 8.00 ± 1.27 | 9.50 ± 0.69 | 36.78 ± 1.33 |
| | Post 2-months | 10.41 ± 1.28 | 9.47 ± 0.86 | 40.76 ± 1.93 |
| NC/LR | Day 0 | 11.79 ± 1.04 | 10.38 ± 0.52 | 50.13 ± 1.91 |
| | FCNef Day 4 | 10.71 ± 1.18 | 8.79 ± 0.62 | 49.86 ± 1.88 |
| | Post 1-month | 9.68 ± 1.26 | 8.77 ± 0.59 | 48.86 ± 2.01 |
| | Post 2-months | 8.45 ± 1.14 | 8.05 ± 0.56 | 46.65 ± 1.52 |

**Supplementary Table 11. Best-fit LME to predict General Depression on Days 0 and 4.**

*Model: General Depression~ first/last Day + Group + (1|Subject)*

*Main factor variables: General Depression = linear, first/last Day = linear, Group = categorical*

| Variables | Estimate | Lower | Upper | SE | *t* Stat | DF | *p* Value |
|---|---|---|---|---|---|---|---|
| (Intercept) | 13.67 | 10.92 | 16.42 | 1.39 | 9.82 | 132 | <0.001 |
| first/last Day | -1.59 | -2.24 | -0.94 | 0.33 | -4.82 | 132 | <0.001 |
| C/LR | -1.63 | -5.19 | 1.92 | 1.80 | -0.91 | 132 | 0.366 |
| NC/LR | -0.04 | -3.56 | 3.49 | 1.78 | -0.02 | 132 | 0.984 |

**Supplementary Table 12. Best-fit LME to predict Brooding on Days 0 and 4.**

*Model: Brooding ~ first/last Day + Group + (1|Subject)*

*Main factor variables: Brooding = linear, first/last Day = linear, Group = categorical*

| Variables | Estimate | Lower | Upper | SE | *t* Stat | DF | *p* Value |
|---|---|---|---|---|---|---|---|
| (Intercept) | 12.18 | 10.51 | 13.85 | 0.84 | 14.43 | 132 | <0.001 |
| First/last Day | -1.29 | -1.88 | -0.71 | 0.30 | -4.37 | 132 | <0.001 |
| C/LR | 0.72 | -1.25 | 2.68 | 0.99 | 0.72 | 132 | 0.471 |
| NC/LR | -0.65 | -2.60 | 1.29 | 0.98 | -0.67 | 132 | 0.506 |

**Supplementary Table 13. Best-fit LME to predict Anxiety on Days 0 and 4.**

*Model: Anxiety ~ first/last Day + Group + (1|Subject)*

*Main factor variables: Anxiety = linear, first/last Day = linear, Group = categorical*

| Variables | Estimate | Lower | Upper | SE | *t* Stat | DF | *p* Value |
|---|---|---|---|---|---|---|---|
| (Intercept) | 46.39 | 42.00 | 50.79 | 2.22 | 20.87 | 132 | <0.001 |
| First/last Day | -1.71 | -2.84 | -0.57 | 0.57 | -2.98 | 132 | 0.004 |
| C/LR | -4.70 | -10.31 | 0.90 | 2.83 | -1.66 | 132 | 0.100 |
| NC/LR | 5.77 | 0.22 | 11.32 | 2.81 | 2.06 | 132 | 0.042 |

**Supplementary Table 14. Best-fit LME to predict whether General Depression differences from baseline differed over the days following FCNef (the Post-Days).**

*Model: General Depression Post-Change ~Post-Day + Group + (1|Subject)*

*Main factor variables: General Depression Post-Change = linear, Post-Day = linear, Group = categorical*

| Name | Estimate | Lower | Upper | SE | *t* Stat | DF | *p* Value |
|---|---|---|---|---|---|---|---|
| (Intercept) | -2.05 | -3.80 | -0.31 | 0.88 | -2.33 | 163 | 0.021 |
| Post-Day | -0.18 | -0.79 | 0.44 | 0.31 | -0.57 | 163 | 0.570 |
| C/LR | 1.01 | -0.91 | 2.92 | 0.97 | 1.04 | 163 | 0.301 |
| NC/LR | 0.56 | -1.33 | 2.45 | 0.96 | 0.58 | 163 | 0.561 |

**Supplementary Table 15. Best-fit LME to predict whether Brooding differences from baseline differed over the days following FCNef (the Post-Days).**

*Model: Brooding Post-Change ~Post-Day + Group + (1|Subject)*

*Main factor variables: Brooding Post-Change = linear, Post-Day = linear, Group = categorical*

| Name | Estimate | Lower | Upper | SE | *t* Stat | DF | *p* Value |
|---|---|---|---|---|---|---|---|
| (Intercept) | -0.49 | -1.63 | 0.66 | 0.58 | -0.84 | 161 | 0.400 |
| Post-Day | -0.34 | -0.71 | 0.04 | 0.19 | -1.78 | 161 | 0.076 |
| C/LR | -0.61 | -1.93 | 0.72 | 0.67 | -0.91 | 161 | 0.366 |
| NC/LR | -0.66 | -1.96 | 0.65 | 0.66 | -0.99 | 161 | 0.322 |

**Supplementary Table 16. Best-fit LME to predict whether Anxiety differences from baseline differed over the days following FCNef (the Post-Days).**

*Model: Anxiety Post-Change ~Post-Day + Group + (1|Subject)*

*Main factor variables: Anxiety Post-Change = linear, Post-Day = linear, Group = categorical*

| Name | Estimate | Lower | Upper | SE | *t* Stat | DF | *p* Value |
|---|---|---|---|---|---|---|---|
| (Intercept) | -1.73 | -4.85 | 1.40 | 1.58 | -1.09 | 161 | 0.278 |
| Post-Day | 0.37 | -0.64 | 1.37 | 0.51 | 0.73 | 161 | 0.469 |
| C/LR | -1.06 | -4.70 | 2.59 | 1.84 | -0.57 | 161 | 0.568 |
| NC/LR | -0.41 | -3.99 | 3.18 | 1.82 | -0.22 | 161 | 0.823 |

**Supplementary Table 17. Almost best-fit LME to predict whether Anxiety differences from baseline differed over the days following FCNef (the Post-Days).**

*Model: Anxiety Post-Change ~Post-Day * Group + (1|Subject)*

*Main factor variables: Anxiety Post-Change = linear, Post-Day = linear, Group = categorical*

| Name | Estimate | Lower | Upper | SE | *t* Stat | DF | *p* Value |
|---|---|---|---|---|---|---|---|
| (Intercept) | -1.52 | -5.79 | 2.75 | 2.16 | -0.70 | 159 | 0.483 |
| Post-Day | 0.25 | -1.86 | 2.35 | 1.07 | 0.23 | 159 | 0.818 |
| C/LR | -3.76 | -9.51 | 1.99 | 2.91 | -1.29 | 159 | 0.198 |
| NC/LR | 1.52 | -4.15 | 7.18 | 2.87 | 0.53 | 159 | 0.598 |
| Post-Day:C/LR | 1.48 | -1.18 | 4.14 | 1.35 | 1.10 | 159 | 0.274 |
| Post-Day:NC/LR | -1.00 | -3.60 | 1.60 | 1.32 | -0.76 | 159 | 0.451 |

**Supplementary Table 18: Average ± standard deviation of rs-FCs on different days of measurement for the three groups**.

| | Day 0 | FCNef Day 1 | FCNef Day 2 | FCNef Day 3 | FCNef Day 4 | Post 1-month | Post 2-months |
|---|---|---|---|---|---|---|---|
| C/HR | 0.039 ± 0.242 | -0.029 ± 0.224 | -0.087 ± 0.190 | -0.041 ± 0.261 | -0.072 ± 0.231 | -0.032 ± 0.238 | -0.067 ± 0.130 |
| C/LR | -0.042 ± 0.209 | -0.165 ± 0.216 | -0.216 ± 0.188 | -0.165 ± 0.219 | -0.110 ± 0.238 | -0.102 ± 0.205 | -0.097 ± 0.164 |
| NC/LR | -0.118 ± 0.279 | -0.109 ± 0.265 | -0.058 ± 0.263 | -0.047 ±0.227 | -0.090 ± 0.215 | -0.045 ± 0.232 | -0.064 ±0.270 |

**Supplementary Table 19. Best-fit LME to predict rs-FC on Days 0 and 4.**

*Model: rs-FC ~FCNef Day + Group + (1|Subject)*

*Main factor variables: rs-FC = linear, FCNef Day = linear, Group = categorical*

| Name | Estimate | Lower | Upper | SE | *t* Stat | DF | *p* Value |
|---|---|---|---|---|---|---|---|
| (Intercept) | 0.05 | -0.06 | 0.17 | 0.06 | 0.90 | 131 | 0.371 |
| FCNef Day | -0.05 | -0.10 | 0.01 | 0.03 | -1.69 | 131 | 0.094 |
| C/LR | -0.06 | -0.19 | 0.06 | 0.06 | -1.00 | 131 | 0.321 |
| NC/LR | -0.09 | -0.21 | 0.03 | 0.06 | -1.47 | 131 | 0.144 |

**Supplementary Table 20. Almost best-fit LME to predict rs-FC on Days 0 and 4**

*Model: rs-FC ~FCNef Day * Group + (1|Subject)*

*Main factor variables: rs-FC = linear, FCNef Day = linear, Group = categorical*

| Name | Estimate | Lower | Upper | SE | *t* Stat | DF | *p* Value |
|---|---|---|---|---|---|---|---|
| (Intercept) | 0.15 | -0.02 | 0.31 | 0.08 | 1.76 | 129 | 0.081 |
| FCNef Day | -0.11 | -0.20 | -0.01 | 0.05 | -2.27 | 129 | 0.025 |
| C/LR | -0.12 | -0.35 | 0.10 | 0.11 | -1.06 | 129 | 0.290 |
| NC/LR | -0.29 | -0.52 | -0.07 | 0.11 | -2.58 | 129 | 0.011 |
| FCNef Day:C/LR | 0.04 | -0.09 | 0.17 | 0.06 | 0.63 | 129 | 0.532 |
| FCNef Day:NC/LR | 0.14 | 0.01 | 0.26 | 0.06 | 2.12 | 129 | 0.036 |

**Supplementary Table 21. Best-fit LME to predict rs-FC Post-Change.**

*Model: rs-FC Post-Change ~ Post-Day + Group + (1|Subject)*

*Main factor variables: rs-FC Post-Change = linear, Post-Day = linear, Group = categorical*

| Name | Estimate | Lower | Upper | SE | *t* Stat | DF | *p* Value |
|---|---|---|---|---|---|---|---|
| (Intercept) | -0.12 | -0.22 | -0.02 | 0.05 | -2.43 | 153 | 0.016 |
| Post-Day | 0.01 | -0.02 | 0.04 | 0.02 | 0.40 | 153 | 0.691 |
| C/LR | 0.05 | -0.07 | 0.16 | 0.06 | 0.85 | 153 | 0.395 |
| NC/LR | 0.16 | 0.04 | 0.27 | 0.06 | 2.76 | 153 | 0.007 |

**Supplementary Table 22. Best-fit LME to predict General Depression Change (Day 4 - Day 0) using rs-FC Change (Day 4 - Day 0).**

*Model: General Depression Change ~ rs-FC Change + Group + (1|Subject)*

*Main factor variables: General Depression Change = linear, rs-FC Change = linear, Group = categorical*

| Name | Estimate | Lower | Upper | SE | *t* Stat | DF | *p* Value |
|---|---|---|---|---|---|---|---|
| (Intercept) | -0.77 | -2.38 | 0.85 | 0.81 | -0.95 | 62 | 0.348 |
| rs-FC Change | 16.70 | 9.40 | 23.99 | 3.65 | 4.57 | 62 | < 0.001 |
| Group | -0.14 | -0.84 | 0.57 | 0.35 | -0.39 | 62 | 0.699 |
| rs-FC Change:Group | -6.83 | -9.89 | -3.76 | 1.53 | -4.45 | 62 | < 0.001 |

**Supplementary Table 23. Best-fit LME to predict Brooding Change (Day 4 - Day 0) using rs-FC Change (Day 4 - Day 0).**

*Model: Brooding Change ~ rs-FC Change + Group + (1|Subject)*

*Main factor variables: Brooding Change = linear, rs-FC Change = linear, Group = categorical*

| Name | Estimate | Lower | Upper | SE | *t* Stat | DF | *p* Value |
|---|---|---|---|---|---|---|---|
| (Intercept) | -0.14 | -1.60 | 1.33 | 0.73 | -0.19 | 62 | 0.854 |
| rs-FC Change | 1.96 | -0.40 | 4.32 | 1.18 | 1.66 | 62 | 0.102 |
| Group | -0.50 | -1.15 | 0.15 | 0.32 | -1.54 | 62 | 0.128 |

**Supplementary Table 24. Best-fit LME to predict Anxiety Change (Day 4 - Day 0) using rs-FC Change (Day 4 - Day 0).**

*Model: Anxiety Change ~ rs-FC Change + Group + (1|Subject)*

*Main factor variables: Anxiety Change = linear, rs-FC Change = linear, Group = categorical*

| Name | Estimate | Lower | Upper | SE | *t* Stat | DF | *p* Value |
|---|---|---|---|---|---|---|---|
| (Intercept) | -1.14 | -4.45 | 2.18 | 1.66 | -0.69 | 64 | 0.495 |
| rs-FC Change | 1.36 | -4.08 | 6.80 | 2.72 | 0.50 | 64 | 0.619 |
| Group | -0.22 | -1.69 | 1.26 | 0.74 | -0.29 | 64 | 0.772 |

384 # References

385 1. Beck, A.T., Steer, R.A., & Brown, G.K. *Beck Depression Inventory (BDI-II): Manual and*
386 *Questionnaire*. (1996).

387 2. Treynor, W., Gonzalez, R. & Nolen-Hoeksema, S. Rumination Reconsidered: A Psychometric
388 Analysis. *Cognit. Ther. Res.* **27**, 247–259 (2003).

389 3. Hasegawa, A. Translation and initial validation of the Japanese version of the Ruminative Responses
390 Scale. *Psychol. Rep.* **112**, 716–726 (2013).

391 4. Spielberger, C. D., Gorsuch, R. L., Lushene, R. & Vagg, P. R. Manual for the State-Trait Anxiety
392 Inventory; Palo Alto, CA, Ed. *Palo Alto: Spielberger* (1983).

393 5. Taylor, J. E. *et al.* Depressive symptoms reduce when dorsolateral prefrontal cortex-precuneus
394 connectivity normalizes after functional connectivity neurofeedback. *Sci. Rep.* **12**, 2581 (2022).

395 6. Feinberg, D. A. *et al.* Multiplexed echo planar imaging for sub-second whole brain FMRI and fast
396 diffusion imaging. *PLoS One* **5**, e15710 (2010).

397 7. Moeller, S. *et al.* Multiband multislice GE-EPI at 7 tesla, with 16-fold acceleration using partial
398 parallel imaging with application to high spatial and temporal whole-brain fMRI. *Magn. Reson. Med.*
399 **63**, 1144–1153 (2010).

400 8. Xu, J. *et al.* Evaluation of slice accelerations using multiband echo planar imaging at 3 T.
401 *Neuroimage* **83**, 991–1001 (2013).
