## Supplementary material for "Paving the way for precision treatment of psychiatric symptoms with functional connectivity neurofeedback": MS Word document of manuscript with no figures or tables

Short Title: FCNef for Precision Psychiatry

Corresponding Author: Jessica Elizabeth Taylor

Keywords: Precision psychiatry, real-time fMRI neurofeedback, Major Depressive Disorder, Rumination

### Abstract

***Introduction***: Major depressive disorder (MDD) remains challenging to treat, with many patients failing to respond adequately to existing therapies. Patients with MDD have heterogeneous subsets of symptoms with differing underlying neural aberrations. Treatment response may improve if treatments become more individualised. We recently showed preliminary evidence that normalisation of a neural network and a corresponding reduction in related symptoms can be achieved using a Brain Machine Interface (BMI) called real-time fMRI functional connectivity neurofeedback (FCNef). However, the robustness of this effect, and the best FCNef parameters for optimising therapeutic outcomes remained unknown. ***Methods:*** We ran additional participants, with a final dataset of N = 68, in our FCNef protocol. Functional connectivity between the dorsolateral prefrontal cortex (DLPFC) and posterior cingulate cortex/precuneus (PCC) was targeted with the goal of reducing brooding rumination symptoms. Core FCNef parameters (experimental schedule and reward) were manipulated between participants. ***Results:*** We replicated findings that normalisation of DLPFC-PCC connectivity with FCNef correlates significantly with reductions in brooding rumination, but not with changes in anxiety, which is associated with different neural circuits. The difference between these correlations was significant, highlighting the precision of this effect. Finally, we found that successful DLPFC-PCC normalisation and corresponding changes in brooding rumination depended on specific FCNef parameters. The most effective protocol involved consecutive training days with greater external reward. ***Conclusions***: These results highlight the potential of FCNef for precision medicine in psychiatry and underscore the importance of optimising parameters to enhance feasibility of BMI-based clinical interventions.

### 1. Introduction

The World Health Organization estimates that major depressive disorders will become the top cause of global disease burden by 2030 [[1]](https://paperpile.com/c/6QyoMw/dV5cV). Of patients who do receive treatment, an estimated 30-50% do not respond fully [[2,3]](https://paperpile.com/c/6QyoMw/eDbvw+EvOuZ). Patients with the same clinical diagnosis can have heterogeneous subsets of symptoms that relate to different underlying neural mechanisms [[4]](https://paperpile.com/c/6QyoMw/DDhMd). Nonetheless, they usually receive relatively homogenous treatment. For example, all or most clinical practice guidelines recommend selective serotonin reuptake inhibitors as first-line treatment for depression [[5]](https://paperpile.com/c/6QyoMw/CY0PX). To improve response rates, individual differences clearly need to be considered. Future treatment may become more individualised using Brain-Machine Interfaces (BMIs) to identify and target underlying patient neural aberrations. However, medical regulatory approval presents serious challenges to this [[6]](https://paperpile.com/c/6QyoMw/fO5wj). To date, only a handful of BMIs have received approval from local medical regulatory agencies for human trials [[7–9]](https://paperpile.com/c/6QyoMw/Zdu0m+wRjA3+ihkXR), and even fewer have received full market authorisation [[10,11]](https://paperpile.com/c/6QyoMw/OrKaJ+uhLCN). A key step toward approval of a given BMI technique by regulatory agencies is demonstrating the optimality of chosen parameters. Here, we extend previous results to better examine the robustness of a promising form of BMI called Functional-Connectivity Neurofeedback (FCNef) [[12]](https://paperpile.com/c/6QyoMw/yRcbH), and to systematically investigate a specific set of its parameters.

FCNef is a functional magnetic resonance imaging (fMRI) neurofeedback technique in which participants receive real-time feedback about the current state of functional connectivity between targeted brain regions (measured as the correlation between time-courses of BOLD activity from these regions). This feedback is used to train participants to make a targeted functional connection more positive or negative, a result that has been demonstrated in multiple studies [[12–19]](https://paperpile.com/c/6QyoMw/Cnpos+gA1c6+5GToE+jFrjC+yRcbH+Y0NB6+YrOeS+2mwGP). Showing promise for precision medicine, recent FCNef studies have reported precise correspondence between normalisation of functional connections and reductions in specifically related symptoms [[12,17,18,20]](https://paperpile.com/c/6QyoMw/q67zd+Y0NB6+yRcbH+YrOeS). Consistent with these results, we previously found that normalisation of functional connectivity between the dorsolateral prefrontal cortex (DLPFC) and the precuneus/posterior cingulate cortex (PCC) that occurred with FCNef was related to reductions in brooding rumination, but not anxiety symptoms. Importantly, these effects persisted at least 1-2 months after FCNef [[12]](https://paperpile.com/c/6QyoMw/yRcbH). Nonetheless these results were reported from only a small sample. We have since continued to collect data and here we examine the robustness of these effects by testing for their replication in the newly collected data. Furthermore, because combining the new and old data provides sufficient statistical power to properly compare correlation coefficients, for the first time, we are able to directly examine the specificity of this effect.

Given its potential use as a medical tool, FCNef should maximise health outcomes while minimising patient burden. Here, we operationalised FCNef success as the normalisation of DLPFC/PCC functional connectivity and a related reduction in brooding (but not anxiety) symptoms. We sought to find parameters that would best enhance this, while also keeping participant fatigue to a minimum (because preliminary testing in a clinical sample caused fatigue-related drop-out of one out of six patients [[21]](https://paperpile.com/c/6QyoMw/37SBk)). To accomplish our objective, we focused on the following parameters: (1) Reward schedule: During real-time neurofeedback tasks, feedback has conventionally been provided simply as scores that reflect how similar the induced brain activity is to the target brain activity [[22–24]](https://paperpile.com/c/6QyoMw/B1Jbv+y4PXE+dLPsj). However, recent evidence suggests that target neural activity may be better reinforced during neurofeedback when external reward, such as money, is also used [[25]](https://paperpile.com/c/6QyoMw/bwOnQ). Here, we manipulated how bonus money was assigned to feedback scores so that different groups of participants could earn less/more overall external reward. (2) Experimental schedule. Most FCNef studies [[15]](https://paperpile.com/c/6QyoMw/5GToE), including our own [[12,19]](https://paperpile.com/c/6QyoMw/2mwGP+yRcbH), have required participants to come in for multiple consecutive days of experimentation. This can be exhausting and requires motivation and organisation skills that can be diminished in psychiatric disorders [[26]](https://paperpile.com/c/6QyoMw/1dM5j). Therefore, we tested whether a more flexible schedule, over non-consecutive days, could yield similar results to the consecutive training schedule.

Details of the symptom questionnaires can be found in the Supplementary Methods, but overall general depressive symptoms were measured with the BDI [[27]](https://paperpile.com/c/6QyoMw/jcpwZ). Brooding rumination symptoms were measured with a subscale of the Rumination Response Scale (RRS) [[28,29]](https://paperpile.com/c/6QyoMw/iRFSQ+EaxOs), and trait anxiety symptoms were measured with the trait anxiety subscale of the State-Trait Anxiety Inventory (STAI-Y2) [[30]](https://paperpile.com/c/6QyoMw/NM8zH). As can be seen in Figure 1, because there would be limited clinical meaning, these symptom questionnaire scores were not measured on all days of the main experiment. Instead, they were only measured on the first day (Day 0) and last day (Day 4) of the main experiment and during one- and two-month follow-up tests.

#### Conflict of interest statement

MK is an inventor of patents related to functional connectivity neurofeedback. The original assignee of the patents is ATR, with which the authors are affiliated. We have no other conflicts of interest.

#### Funding Sources

This research was supported by the Japan Agency for Medical Research and Development (AMED) under Grant Numbers JP18dm0307008, JP21dm0307102 and JP17dm0107044, by the Japan Society for the Promotion of Science (JSPS) KAKENHI under Grant Number 24K22822, and by the Innovative Science and Technology Initiative under Security Grant Number JPJ004596, ATLA, Japan.

#### Author Contributions

Jessica Elizabeth Taylor, Tomokazu Motegi, Takashi Yamada and Mitsuo Kawato designed experiments; Jessica Elizabeth Taylor, Taiki Oka, Misa Murakami and Tomokazu Motegi acquired data; Jessica Elizabeth Taylor analysed data; Jessica Elizabeth Taylor prepared the original draft; Taiki Oka, Misa Murakami, Tomokazu Motegi, Takashi Yamada, Takahiko Kawashima, Yuko Kobayashi, Yujiro Yoshihara, Jun Miyata, Toshiya Murai, Mitsuo Kawato, and Aurelio Cortese reviewed and edited the manuscript. All authors gave final approval for submission and agreed to take responsibility for the manuscript.

#### Data accessibility

Data and code supporting this study's findings will be publicly available on our GitHub at publication.

### Figure Legends

#### **Figure 1. Experimental procedure and example FCNef trial.**

***a. The order of events on each day of experimentation.*** *Questionnaires = the Beck Depression Inventory-II* [*[27]*](https://paperpile.com/c/6QyoMw/jcpwZ)*, the Rumination Response Scale* [*[28]*](https://paperpile.com/c/6QyoMw/iRFSQ)*, and the State-Trait Anxiety Inventory* [*[30]*](https://paperpile.com/c/6QyoMw/NM8zH)*. Anatomical = T1-weighted structural MRI. N-back = a well-known executive control task* [*[54]*](https://paperpile.com/c/6QyoMw/lHjSx)*, used here as a functional localiser*[*[12]*](https://paperpile.com/c/6QyoMw/yRcbH)*.* ***b. An example FCNef trial.*** *During the rest period, participants were to simply relax. During the induction period, they were asked to “somehow” manipulate their brain activity to get the best possible feedback. Participants were told that different strategies of brain activity manipulation might work for different people. Unbeknown to participants (nothing changed on screen), there was a 2s calculation period at the end of the induction period. During FCNef, DLPFC-PCC connectivity (from the induction period) was calculated during the calculation period and this determined the feedback presented during the feedback period. During SHAM, however, feedback was just random. Feedback was presented on screen as a green circle and participants had been clearly instructed that the larger this was, the more monetary reward they would receive on that trial. During FCNef, they were instructed to try to make the green circle bigger than the red circle that was also presented on screen. The circumference of this red circle represented the participant's baseline DLPFC-PCC connectivity (the average from SHAM). During SHAM, there was no red circle and participants were simply instructed to try to make the green circle as big as possible. Modified with permission from Taylor et al. (2022)*[*[12]*](https://paperpile.com/c/6QyoMw/yRcbH)*.*
